# Early Subtypes and Progressions of Progressive Supranuclear Palsy: A Data-Driven Brain Bank Study

**DOI:** 10.1101/2025.07.04.25330863

**Authors:** Daisuke Ono, Hiroaki Sekiya, Nikhil B. Ghayal, Alexia R. Maier, Shanu F. Roemer, Ryan J. Uitti, Irene Litvan, Keith A. Josephs, Zbigniew K. Wszolek, Dennis W. Dickson

## Abstract

**Background:** Progressive supranuclear palsy (PSP) is typically characterized by vertical supranuclear gaze palsy and early falls, referred to as Richardson’s syndrome (PSP-RS). Other presentations include postural instability (PSP-PI), parkinsonism (PSP-P), speech/language disorder (PSP-SL), frontal presentation (PSP-F), ocular motor dysfunction (PSP-OM), and corticobasal syndrome (PSP-CBS). Differences across the early presentations and in their subsequent progression have yet to be elucidated.

**Objective:** This study aimed to characterize early PSP subtypes and their subsequent progressions using a large postmortem dataset.

**Methods:** An automated pipeline incorporating fine-tuned Chat Generative Pre-trained Transformer (ChatGPT) was developed. The pipeline collected 195 clinical features with onset information from autopsy-confirmed PSP cases without significant neurodegenerative co-pathologies.

**Results:** A structured clinicopathologic dataset from 588 patients was analyzed. After distilling results with unsupervised clustering, a decision tree model was developed. With five clinical manifestations: frontal presentation, postural instability, ocular motor dysfunction, speech/language disorder, and parkinsonism, this mutually exclusive algorithm identified seven subtypes: PSP-PF (postural and frontal dysfunction), PSP-RS, PSP-PI, PSP-P, PSP-SL, PSP-F, and PSP-OM. PSP-PF showed rapid progression, the shortest median disease duration (six years), and high tau burden in cortical and subcortical regions. In PSP-F, frontal presentation preceded other symptoms by four years, with a nine-year disease duration—second longest after PSP-P (10 years). PSP-CBS was not identified as an independent subtype.

**Conclusions:** This data-driven study identified a novel, aggressive PSP phenotype characterized by early postural and frontal dysfunction. Early subtyping utilizing the decision tree model would help clinicians estimate progression and facilitate early patient recruitment for clinical trials.

## Introduction

Progressive Supranuclear Palsy (PSP) was originally described as a progressive brain disease with vertical supranuclear ophthalmoplegia, recurrent falls, pseudobulbar palsy, axial rigidity, and dementia.[1, 2] It is now defined neuropathologically as a four-repeat tauopathy, marked by neuronal and glial tau pathology, including globose neurofibrillary tangles, tufted astrocytes, and coiled bodies.[3–5] Previous clinical criteria for PSP emphasized the presence of vertical supranuclear gaze palsy and early falls within three years of onset, referred to as Richardson’s syndrome (PSP-RS). [6] Thereafter, other phenotypes, where gaze palsy and falls are not predominant in the early stages, have also been reported, such as parkinsonism-predominant (PSP-P)[7] and progressive gait freezing (PSP-PGF).[8]

To expand diagnoses of PSP to include these non-RS subtypes, the Movement Disorder Society’s clinical diagnostic criteria for PSP (MDS-PSP criteria) defined eight clinical subtypes: PSP-RS, PSP-P, PSP-PGF, ocular motor dysfunction (PSP-OM), postural instability (PSP-PI), frontal presentation (PSP-F), speech/language disorder (PSP-SL), and corticobasal syndrome (PSP-CBS).[9] These subtypes are determined by combination patterns of four functional domains, each with three levels of diagnostic certainty. Although this approach improved sensitivity to non-RS subtypes, the criteria were not mutually exclusive, often resulting in multiple diagnoses. The Multiple Allocations eXtinction (MAX) rule was subsequently proposed to prioritize diagnoses based on certainty, temporal order, and phenotypic hierarchy.[10] While this rule reduced the number of diagnoses, it disproportionately favored PSP-RS, with diagnoses frequently shifting to PSP-RS as the disease progressed, potentially underestimating phenotypic diversity.[11–13]

Early signs and symptoms have been reported to characterize subsequent progression and prognosis in neurodegenerative disease. [7, 8, 14–16] Respondek et al. implemented these findings in the development of criteria for PSP phenotypes based on clinical presentations within two years of onset.[14] The Respondek criteria successfully classified 100 autopsy-conformed PSP cases into early subtypes with different syndromes and prognoses, although 13% of the cases were unclassified after the clinicians’ review due to various combinations of symptoms. An objective, mutually exclusive subtyping algorithm that evaluates the early clinical features remains to be established.

To investigate disease progression, symptoms need to be abstracted with their onset information, which requires substantial effort and has been a significant obstacle to large-scale analysis. Recently, large language models (LLMs) have been explored for automating clinical data collection.[17–21] LLMs have successfully performed relatively simple tasks, such as extracting scores and dates from neuropsychiatric tests,[17] clinical presentations,[18, 21, 22] or pathological findings.[19, 20] However, no study to date has successfully applied LLMs to abstract the presence of symptoms with its onset, despite their importance in understanding neurodegenerative diseases. This is partially due to current performance constraints of LLMs with respect to complex tasks and a lack of datasets that have reliable annotations.

This study aimed to define and characterize early clinical subtypes of PSP in an objective and mutually exclusive manner using a large autopsy-confirmed dataset. To achieve this, we developed a fully automated pipeline incorporating a fine-tuned ChatGPT model at three steps to abstract clinical features with respect to presentation and onset. An unsupervised clustering algorithm using 12 parameters identified six early PSP subtypes, which were used to simplify the classification algorithm. An updated decision tree model successfully classified early PSP subtypes, demonstrating distinct early phenotypes, subsequent progression, and prognoses.

## Methods

### Subjects

This retrospective study was approved by the Mayo Clinic Institutional Review Board (24-006694). Patients with a neuropathologic diagnosis of PSP were identified by searching the Mayo Clinic Brain Bank Florida database from 1998 to 2022 (Fig. 1). Patients with co-pathologies of other significant neurodegenerative diseases, such as Alzheimer’s disease (AD), Lewy body disease (LBD), including incidental Lewy bodies, multiple system atrophy (MSA), frontotemporal lobar degeneration (FTLD), corticobasal degeneration (CBD), or major cerebrovascular disease were excluded. Patients with pathological aging (senile plaques with limited or no tau pathology)[23, 24] or primary age-related tauopathy were not explicitly excluded, as their coexistence is difficult to demonstrate in the presence of PSP. Patients with argyrophilic grain disease (AGD) or limbic-predominant age-related TDP-43 encephalopathy were included to evaluate their contribution to clinical presentation.

**Fig. 1.**
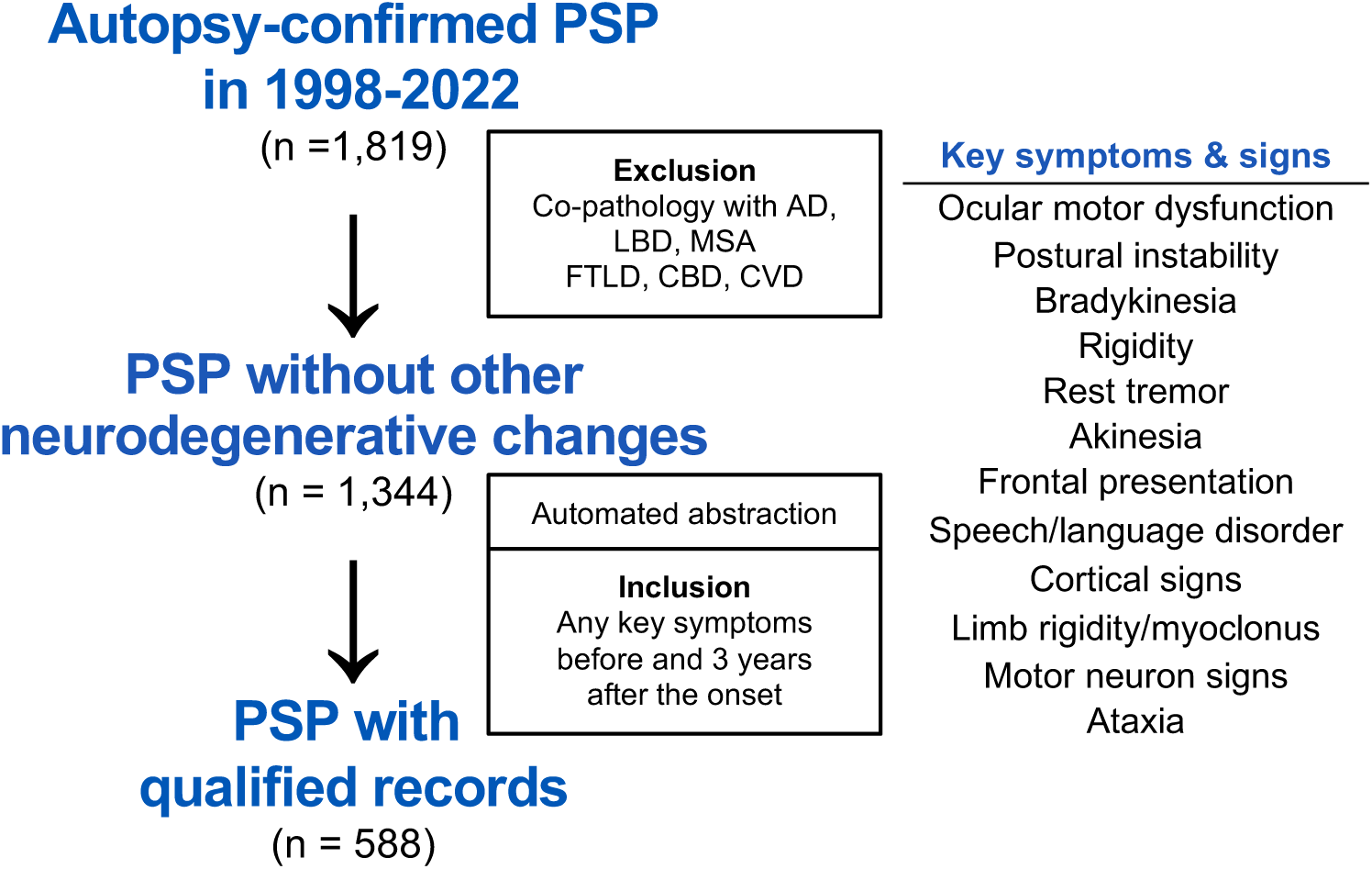
Patients’ selection flow. Among 1,819 autopsy-confirmed patients with progressive supranuclear palsy (PSP), those with co-pathology were excluded, resulting in 1,344 PSP cases without other neurodegenerative changes. After automated abstraction of clinical records, 588 cases with documentation of any of 12 key symptoms and signs at least two different time points—one within three years of disease onset and one after three years— were included for further analysis. Alzheimer’s disease, AD; corticobasal degeneration, CBD; cerebrovascular disease, CVD; frontotemporal lobar degeneration, FTLD; Lewy body disease, LBD; multiple system atrophy, MSA

To capture the broad spectrum of early PSP subtypes, we adopted the following key 12 symptoms and signs, with reference to the subtypes defined by the MDS-PSP criteria[9] and other potential subtypes not defined by it.[25–28] These included ocular motor dysfunction (OM), postural instability (PI, including repeated fall and falling backwards), bradykinesia, rigidity, rest tremor, akinesia, frontal presentation (behavioral or dysexecutive features), speech/language disorder (SL), cortical signs (apraxia, cortical sensory deficit, simultanagnosia, or alien limb phenomena), limb rigidity/limb myoclonus (corresponding to “movement disorder signs” of CBS in the MDS-PSP criteria[9]), upper or lower motor neuron signs, and ataxia. Patients with any descriptions related to these symptoms in the clinicians’ notes and test results at least two different time points—one within three years of disease onset and one after three years— were included for further analysis.

### Neuropathologic evaluation

Neuropathologic evaluations were performed by DWD. The pathologic diagnoses of PSP and concomitant co-pathologies were made according to previous publications.[3, 4, 29–35] Braak Neurofibrillary tangle (NFT) stage (0-VI) and Thal amyloid phase (0-5) were determined based on the distribution and counts of NFTs and senile plaques on thioflavin S fluorescence, respectively.[36, 37] Tau pathology scores were assessed as previously described.[38, 39] Briefly, the severity of four tau pathologies: NFTs/pretangles, coiled bodies, tufted astrocytes and tau threads, were semi-quantitatively scored on a four-point scale (0 = absent, 1 = mild, 2 = moderate, 3 = severe) in 21 brain regions with CP-13 immunostained slides. For each region, the scores of these four pathologies (each ranging from 0 to 3) were summed to yield a total tau score (0–12), with reference to previous reports. [13, 39, 40]

### Clinical record abstraction pipeline

A previously published program, which automatically determined the presence of symptoms,[22] was further developed to include additional functions. The updated pipeline consists of three steps: 1) indexing patients’ documents, 2) determining the presence of symptoms, and 3) identifying the onset of symptoms (Supplementary Fig. 1 and Supplementary Tables 1-3). Detailed in the Supplementary Methods, de-identified excerpts from patient records were used at each step to fine-tune a large language model, GPT-4o, with human labels as the gold standard.

### Clustering analysis

Uniform Manifold Approximation and Projection (UMAP) clustering analysis with a Euclidean metric was performed using a python library UMAP-learn 0.5.6 with 15 neighbors, a minimum distance of 0.1, and 2 components. The presence or absence of 12 key symptoms within 3 years of disease onset was used as input, with unavailable data considered as absence.

### Simplified decision tree model

To simplify the subtyping algorithm by the clustering analysis, we applied a decision tree model using scikit-learn 1.5.2. The model used the Gini impurity criterion, with the best splitter strategy, a maximum tree depth of 4, a minimum of 2 samples required to split an internal node, and at least 1 sample required at each leaf node. The model was trained with the same 12 parameters for the clustering analysis. We applied 10-fold cross-validation and evaluated performance using accuracy and area under the curve (AUC). The contribution of each of the 12 parameters to performance was then examined by excluding them one at a time. The decision tree model was simplified by removing non-contributory parameters. The final model, trained on the full dataset using only the significant parameters, was subsequently refined based on domain knowledge, as described in the Results section.

### Survival analysis

We fit multivariable Cox proportional-hazards models incorporating age, sex, and subtype information. Survival curves were generated for a representative patient profile with mean age and male sex. To evaluate the contribution of early PSP subtyping to prognostic prediction, concordance indices (C-indices) were compared between models with and without subtype variables, adjusting for age and sex.

### Statistical analysis

All statistical analyses were performed using scikit-learn 1.5.2, SciPy 1.9.3, and other related Python libraries. Statistical differences in categorical variables between subtypes were evaluated using the chi-square test, followed by the pairwise chi-square test with Holm’s correction for those with significant differences. Continuous values were considered nonparametric and are presented as the median [interquartile range] (number of cases), unless otherwise noted. Statistical differences in continuous variables between subtypes were evaluated using the Kruskal-Wallis test, followed by the pairwise Steel-Dwass test as a post-hoc analysis. *P* value < 0.05 was considered statistically significant.

## Results

### Patients’ selection and characteristics

Among 9,640 brain donors in the Mayo Clinic Brain Bank Florida, 1,819 autopsy-confirmed PSP patients were identified. Patients with significant co-pathologies were excluded: cerebrovascular disease (n = 206), AD (n = 181), LBD (n = 126), CBD (n = 9), MSA (n = 4), ALS (n = 2), FTLD (n = 1), and Pick’s disease (n = 1); numbers are not mutually exclusive, with 1,344 patients undergoing structured clinical record abstraction. Among these, 666 had concomitant amyloid pathology (Thal amyloid phase ≥ 1), 316 had AGD, and 67 had TDP-43 pathology in the amygdala. PSP cases without any of these co-pathologies totaled 383 (21%). After applying the pipeline to 53,527 pages of medical records from these 1,344 patients, 588 patients were identified as meeting the inclusion criteria, with 57% being male, 96% being Caucasian, and the median age of death being 74 years (Fig. 1 and Table 1). Of these, 583 patients (99.1%) had been evaluated by neurologists; 283 brains (48.1%) were sent from specialist clinics or hospitals, while the remaining 305 brains (51.9%) came from regional pathology laboratories and medical examiner offices. Most patients were clinically diagnosed with PSP (87%), while others were diagnosed with parkinsonian disorders, such as corticobasal syndrome (12%), Parkinson’s disease (4%) and multiple system atrophy (3%), including cases with multiple diagnoses.

**Table 1.**
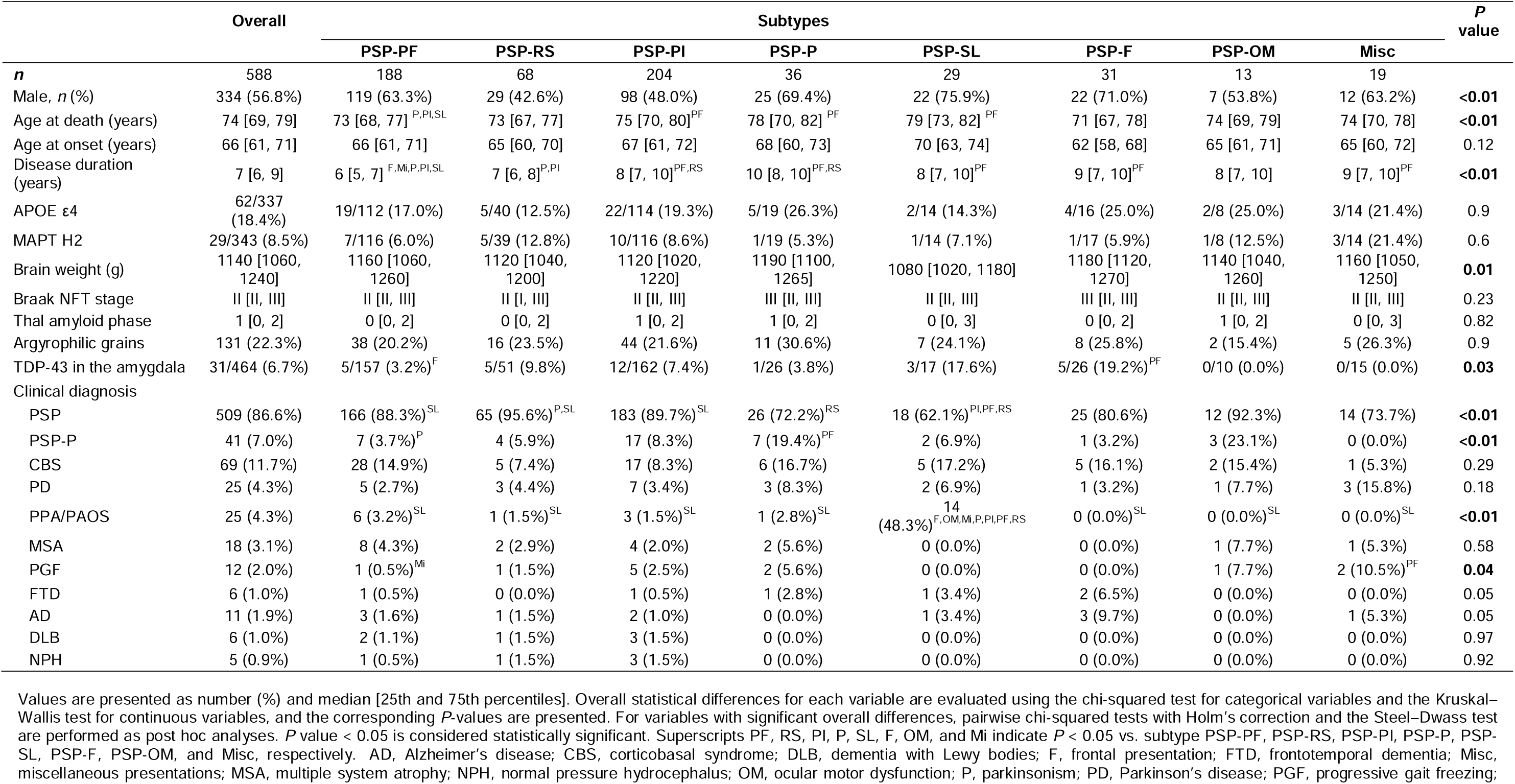
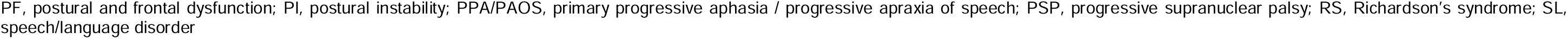
Patients characteristics between subtypes.

|  | Overall | Subtypes |  |  |  |  |  |  |  | P value |
| --- | --- | --- | --- | --- | --- | --- | --- | --- | --- | --- |
|  |  | PSP-PF | PSP-RS | PSP-PI | PSP-P | PSP-SL | PSP-F | PSP-OM | Misc |  |
| **n** | 588 | 188 | 68 | 204 | 36 | 29 | 31 | 13 | 19 |  |
| Male, n (%) | 334 (56.8%) | 119 (63.3%) | 29 (42.6%) | 98 (48.0%) | 25 (69.4%) | 22 (75.9%) | 22 (71.0%) | 7 (53.8%) | 12 (63.2%) | **<0.01** |
| Age at death (years) | 74 [69, 79] | 73 [68, 77]<sup>P,PI,SL</sup> | 73 [67, 77] | 75 [70, 80]<sup>PF</sup> | 78 [70, 82]<sup>PF</sup> | 79 [73, 82]<sup>PF</sup> | 71 [67, 78] | 74 [69, 79] | 74 [70, 78] | **<0.01** |
| Age at onset (years) | 66 [61, 71] | 66 [61, 71] | 65 [60, 70] | 67 [61, 72] | 68 [60, 73] | 70 [63, 74] | 62 [58, 68] | 65 [61, 71] | 65 [60, 72] | 0.12 |
| Disease duration (years) | 7 [6, 9] | 6 [5, 7]<sup>F,Mi,P,PI,SL</sup> | 7 [6, 8]<sup>P,PI</sup> | 8 [7, 10]<sup>PF,RS</sup> | 10 [8, 10]<sup>PF,RS</sup> | 8 [7, 10]<sup>PF</sup> | 9 [7, 10]<sup>PF</sup> | 8 [7, 10] | 9 [7, 10]<sup>PF</sup> | **<0.01** |
| APOE ε4 | 62/337 (18.4%) | 19/112 (17.0%) | 5/40 (12.5%) | 22/114 (19.3%) | 5/19 (26.3%) | 2/14 (14.3%) | 4/16 (25.0%) | 2/8 (25.0%) | 3/14 (21.4%) | 0.9 |
| MAPT H2 | 29/343 (8.5%) | 7/116 (6.0%) | 5/39 (12.8%) | 10/116 (8.6%) | 1/19 (5.3%) | 1/14 (7.1%) | 1/17 (5.9%) | 1/8 (12.5%) | 3/14 (21.4%) | 0.6 |
| Brain weight (g) | 1140 [1060, 1240] | 1160 [1060, 1260] | 1120 [1040, 1200] | 1120 [1020, 1220] | 1190 [1100, 1265] | 1080 [1020, 1180] | 1180 [1120, 1270] | 1140 [1040, 1260] | 1160 [1050, 1250] | **0.01** |
| Braak NFT stage | II [II, III] | II [II, III] | II [I, III] | II [II, III] | III [II, III] | II [II, III] | III [II, III] | II [II, III] | II [II, III] | 0.23 |
| Thal amyloid phase | 1 [0, 2] | 0 [0, 2] | 0 [0, 2] | 1 [0, 2] | 1 [0, 2] | 0 [0, 3] | 0 [0, 2] | 1 [0, 2] | 0 [0, 3] | 0.82 |
| Argyrophilic grains | 131 (22.3%) | 38 (20.2%) | 16 (23.5%) | 44 (21.6%) | 11 (30.6%) | 7 (24.1%) | 8 (25.8%) | 2 (15.4%) | 5 (26.3%) | 0.9 |
| TDP-43 in the amygdala | 31/464 (6.7%) | 5/157 (3.2%)<sup>F</sup> | 5/51 (9.8%) | 12/162 (7.4%) | 1/26 (3.8%) | 3/17 (17.6%) | 5/26 (19.2%)<sup>PF</sup> | 0/10 (0.0%) | 0/15 (0.0%) | **0.03** |
| Clinical diagnosis |  |  |  |  |  |  |  |  |  |  |
| PSP | 509 (86.6%) | 166 (88.3%)<sup>SL</sup> | 65 (95.6%)<sup>P,SL</sup> | 183 (89.7%)<sup>SL</sup> | 26 (72.2%)<sup>RS</sup> | 18 (62.1%)<sup>PI,PF,RS</sup> | 25 (80.6%) | 12 (92.3%) | 14 (73.7%) | **<0.01** |
| PSP-P | 41 (7.0%) | 7 (3.7%)<sup>P</sup> | 4 (5.9%) | 17 (8.3%) | 7 (19.4%)<sup>PF</sup> | 2 (6.9%) | 1 (3.2%) | 3 (23.1%) | 0 (0.0%) | **<0.01** |
| CBS | 69 (11.7%) | 28 (14.9%) | 5 (7.4%) | 17 (8.3%) | 6 (16.7%) | 5 (17.2%) | 5 (16.1%) | 2 (15.4%) | 1 (5.3%) | 0.29 |
| PD | 25 (4.3%) | 5 (2.7%) | 3 (4.4%) | 7 (3.4%) | 3 (8.3%) | 2 (6.9%) | 1 (3.2%) | 1 (7.7%) | 3 (15.8%) | 0.18 |
| PPA/PAOS | 25 (4.3%) | 6 (3.2%)<sup>SL</sup> | 1 (1.5%)<sup>SL</sup> | 3 (1.5%)<sup>SL</sup> | 1 (2.8%)<sup>SL</sup> | 14 (48.3%)<sup>F,OM,Mi,P,PI,PF,RS</sup> | 0 (0.0%)<sup>SL</sup> | 0 (0.0%)<sup>SL</sup> | 0 (0.0%)<sup>SL</sup> | **<0.01** |
| MSA | 18 (3.1%) | 8 (4.3%) | 2 (2.9%) | 4 (2.0%) | 2 (5.6%) | 0 (0.0%) | 0 (0.0%) | 1 (7.7%) | 1 (5.3%) | 0.58 |
| PGF | 12 (2.0%) | 1 (0.5%)<sup>Mi</sup> | 1 (1.5%) | 5 (2.5%) | 2 (5.6%) | 0 (0.0%) | 0 (0.0%) | 1 (7.7%) | 2 (10.5%)<sup>PF</sup> | **0.04** |
| FTD | 6 (1.0%) | 1 (0.5%) | 0 (0.0%) | 1 (0.5%) | 1 (2.8%) | 1 (3.4%) | 2 (6.5%) | 0 (0.0%) | 0 (0.0%) | 0.05 |
| AD | 11 (1.9%) | 3 (1.6%) | 1 (1.5%) | 2 (1.0%) | 0 (0.0%) | 1 (3.4%) | 3 (9.7%) | 0 (0.0%) | 1 (5.3%) | 0.05 |
| DLB | 6 (1.0%) | 2 (1.1%) | 1 (1.5%) | 3 (1.5%) | 0 (0.0%) | 0 (0.0%) | 0 (0.0%) | 0 (0.0%) | 0 (0.0%) | 0.97 |
| NPH | 5 (0.9%) | 1 (0.5%) | 1 (1.5%) | 3 (1.5%) | 0 (0.0%) | 0 (0.0%) | 0 (0.0%) | 0 (0.0%) | 0 (0.0%) | 0.92 |

Values are presented as number (%) and median [25th and 75th percentiles]. Overall statistical differences for each variable are evaluated using the chi-squared test for categorical variables and the Kruskal–Wallis test for continuous variables, and the corresponding *P*-values are presented. For variables with significant overall differences, pairwise chi-squared tests with Holm’s correction and the Steel–Dwass test are performed as post hoc analyses. *P* value < 0.05 is considered statistically significant. Superscripts PF, RS, PI, P, SL, F, OM, and Mi indicate *P* < 0.05 vs. subtype PSP-PF, PSP-RS, PSP-PI, PSP-P, PSP-SL, PSP-F, PSP-OM, and Misc, respectively. AD, Alzheimer’s disease; CBS, corticobasal syndrome; DLB, dementia with Lewy bodies; F, frontal presentation; FTD, frontotemporal dementia; Misc, miscellaneous presentations; MSA, multiple system atrophy; NPH, normal pressure hydrocephalus; OM, ocular motor dysfunction; P, parkinsonism; PD, Parkinson’s disease; PGF, progressive gait freezing;
PF, postural and frontal dysfunction; PI, postural instability; PPA/PAOS, primary progressive aphasia / progressive apraxia of speech; PSP, progressive supranuclear palsy; RS, Richardson's syndrome; SL, speech/language disorder

### Unsupervised subtyping with 12 early presentations

We then performed UMAP clustering analysis with the dataset on the presence or absence of 12 key symptoms within 3 years of disease onset, and we identified six distinct clusters, termed subtype (S) 1 to 6 (Fig. 2a). These clusters revealed significant differences in disease duration, which was shortest in S1 (median: 6 years [interquartile range: 5, 8]) and longest in S4 (9 years [7, 10]) and S6 (9 years [8, 11]) (P < 0.01). To assess the prognostic value of the early subtypes, we fit multivariable Cox proportional-hazards models. Compared with S1, subtypes S2–S6 were associated with significantly lower mortality risk (hazard ratios 0.55– 0.37, all P < 0.01; Fig. 2b and Supplementary Table 4). Incorporating early subtypes into the model increased the C-index from 0.554 to 0.658 (Δ = 0.104) after adjusting for age and sex, indicating a substantial improvement in prognostic prediction.

**Fig. 2.**
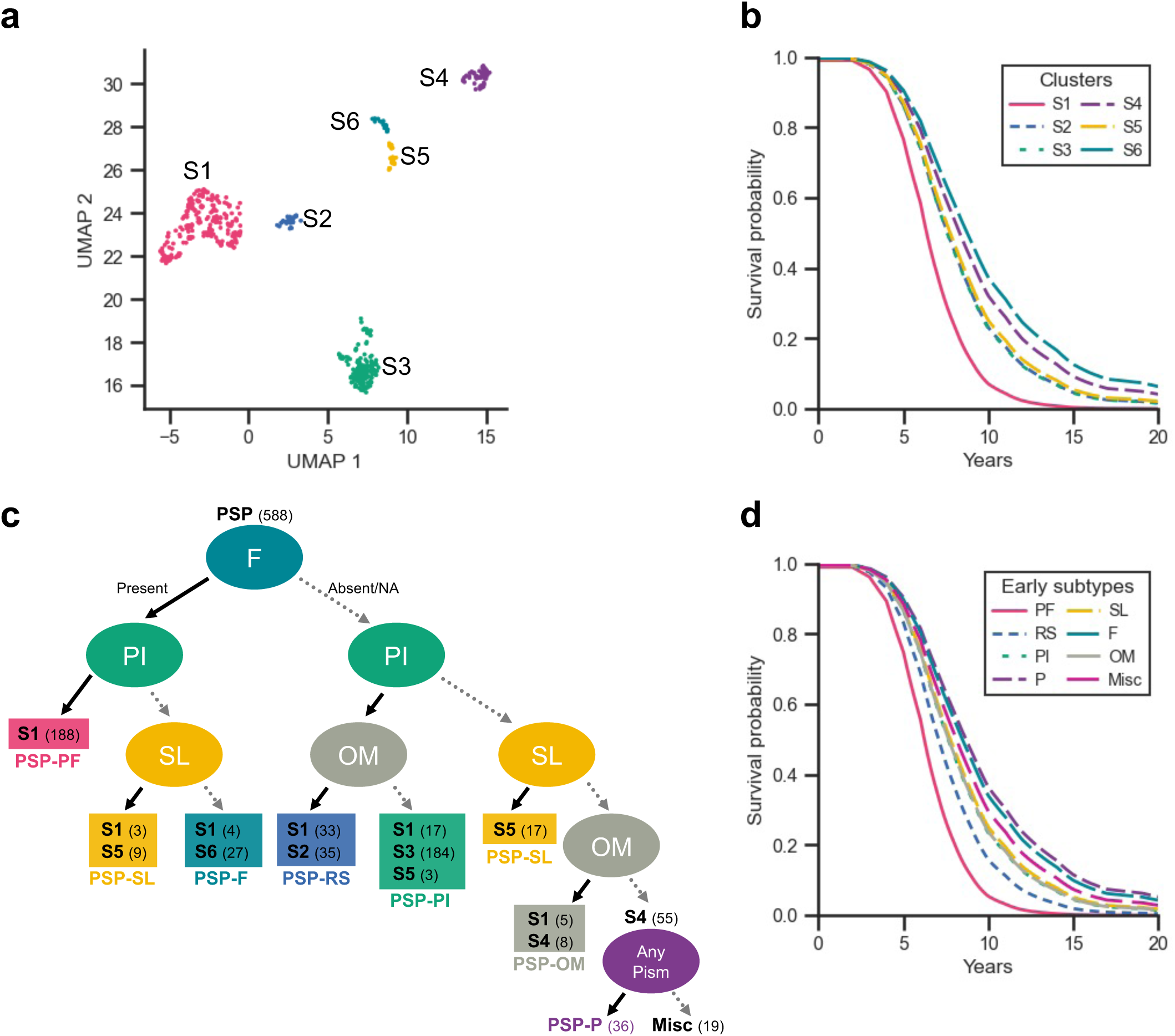
UMAP clustering analysis and early PSP subtypes classified using a simplified decision tree model. (**a**) UMAP, by the presence or absence of 12 key symptoms within 3 years of disease onset, identifies six distinct clusters: subtype (S) 1 to 6. (**b**) Age- and sex-adjusted survival curves between six clusters. **(c)** A decision tree model is optimized to effectively predict the clusters (S1-S6) using a minimal set of clinical features. After adding the leaf of any parkinsonian features to differentiate PSP-P, a simplified decision tree is generated. Solid lines indicate the presence of symptoms, while dashed lines indicate the absence of symptoms or cases where symptoms were not evaluated. Each terminal node displays the subtype(s) along with the number of patients in parentheses. (**d**) Age- and sex-adjusted survival curves between the early PSP subtypes identified using the simplified decision tree model. F, frontal presentation; Misc, miscellaneous presentations; NA, not available; OM, ocular motor dysfunction; P, parkinsonism; PF, postural and frontal dysfunction; PI, postural instability; PSP, progressive supranuclear palsy; RS, Richardson’s syndrome; SL, speech/language disorders; UMAP, uniform manifold approximation and projection

### Simplified decision tree model for early PSP subtyping

To ensure the interpretability and reproducibility of the subtyping in clinical setting, we distilled the UMAP model into a decision tree model with the reduced number of variables, preserving prognostic predictability. We trained a decision tree model to predict S1-S6, examined the effect of each input symptom on model performance, and found that four features—frontal presentation, PI, SL and OM—played critical roles in the subtyping. The simplified decision tree model that used these four parameters successfully reproduced the classification of the UMAP model, achieving an accuracy of 0.878 ± 0.034 and AUC of 0.979 ± 0.014 (mean ± SD). After consulting previous literature, the presence of any parkinsonian features was added to the model, enabling the differentiation of seven distinct subtypes.[7] The resulting seven early PSP subtypes were termed as follows: PSP-PF (postural and frontal dysfunction), PSP-RS, PSP-PI, PSP-P, PSP-SL, PSP-F, and PSP-OM (Fig. 2c). A small number of cases (19/588, 3%) were classified as miscellaneous, including four with any ataxic signs, two with upper neuron signs, one with cortical sign, one with akinesia, and 11 without any positive findings pertaining to the 12 key symptoms.

These early subtypes showed significant differences in disease duration. PSP-PF had the shortest duration of six years [5, 7], while PSP-P had the longest duration of ten years [8, 10]) (P < 0.01, Table 1). Similarly, age- and sex-adjusted survival curves, generated using a multivariable Cox proportional-hazards model, demonstrated lower mortality risks for all other subtypes compared to PSP-PF (hazard ratios 0.63–0.35, all P < 0.01; Fig. 2d and Supplementary Table 5). Incorporating subtype information increased the C-index from 0.554 to 0.654 (Δ = 0.100), which was comparable to the prognostic performance of the UMAP clustering model.

### Clinical characteristics of early PSP subtypes

The clinical presentation and progression of the early PSP subtypes are visualized in Figs. 3 and 4 (the raw data presented in Supplementary Table 6). Patients with PSP-PF presented with PI (half of the patients developed this symptom 0.3 years after disease onset) and frontal presentations (1.6 years). They developed bradykinesia (2.1 years), rigidity (2.5 years), and OM (2.6 years) in the early stage. Patients with PSP-RS initially exhibited PI (0.6 years) and OM (1.8 years) and later developed bradykinesia (2.3 years) and rigidity (3.0 years). Patients in PSP-PI experienced early PI (0.3 years), followed by the development of bradykinesia (4.4 years), rigidity (4.5 years), and OM (4.7 years). The PSP-P, PSP-SL, PSP-F, and PSP-OM subtypes were characterized by early presentations of tremor (1.6 years), SL (0.6 years), frontal presentation (0.3 years), and OM (1.5 years), respectively.

**Fig. 3.**
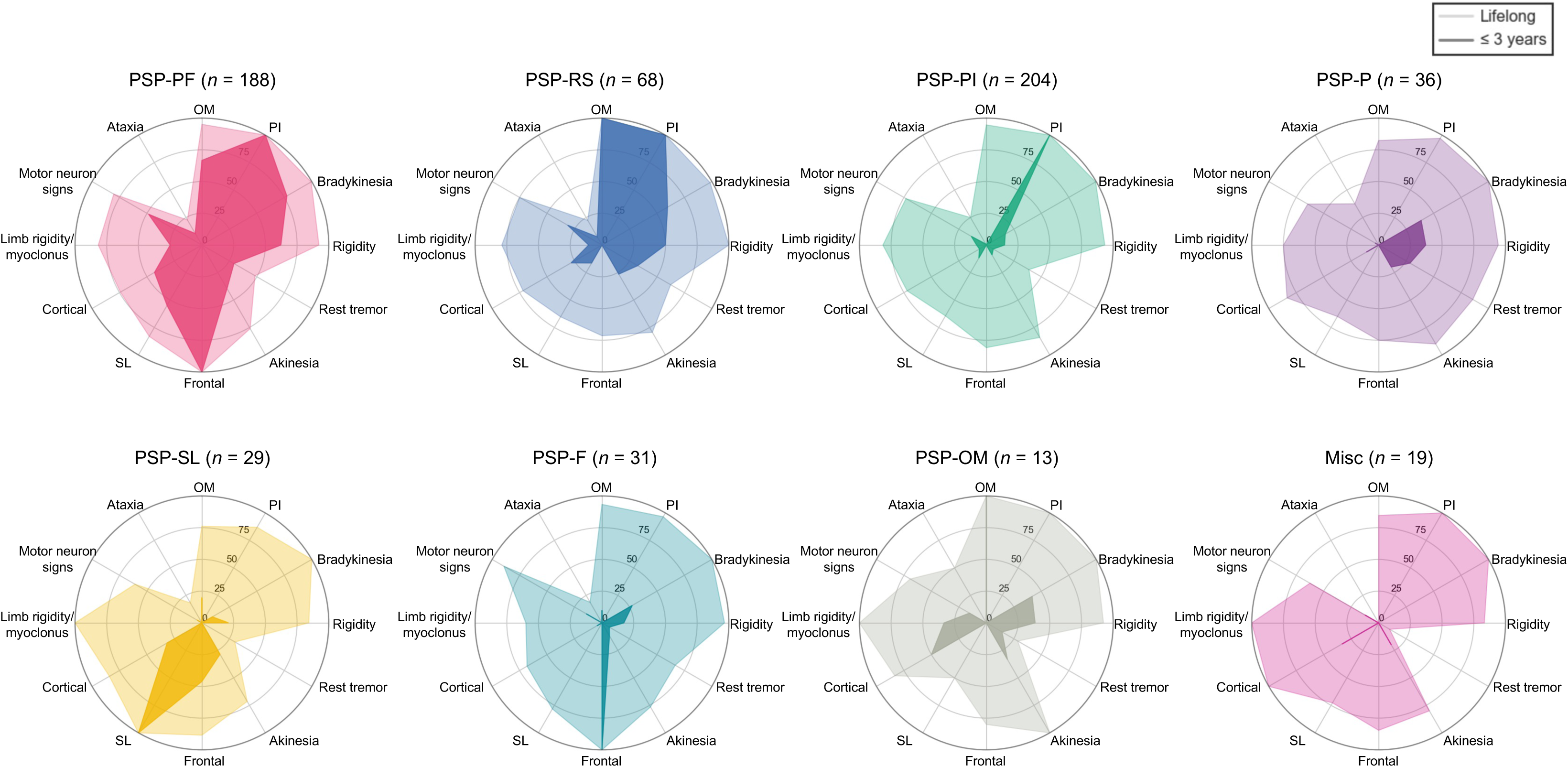
Early and lifelong clinical features of PSP between subtypes. Radar plots show percentages calculated by dividing the number of cases present within three years after onset by the number of cases evaluated at death (dark colour), and by dividing the number of cases present at death by the number of cases evaluated at death (light colour). F, frontal presentation; Misc, miscellaneous presentations; OM, ocular motor dysfunction; P, parkinsonism; PF, postural and frontal dysfunction; PI, postural instability; PSP, progressive supranuclear palsy; RS, Richardson’s syndrome; SL, speech/language disorders

**Fig. 4.**
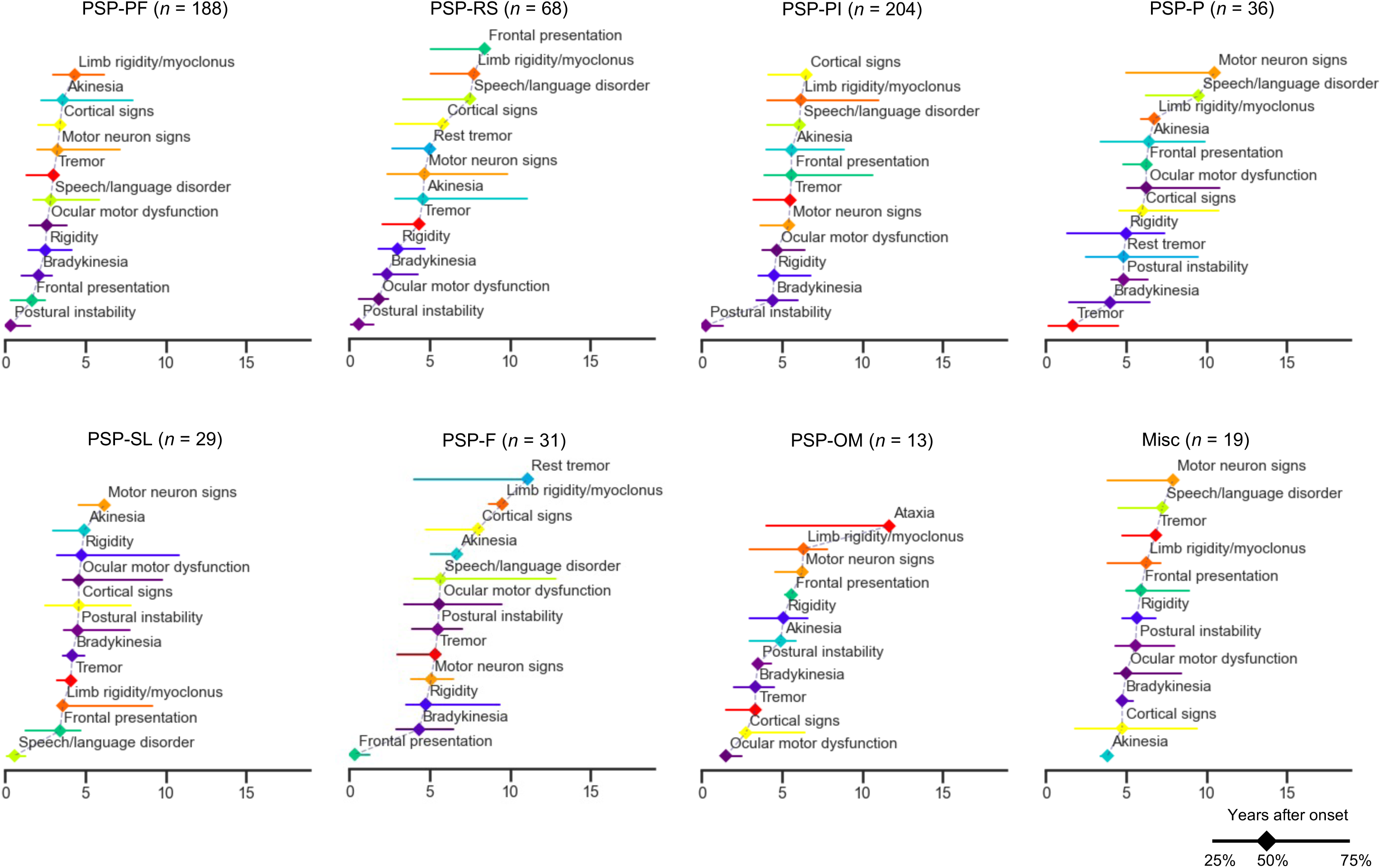
Progression of syndromes in early PSP subtypes. Clinical progression patterns between PSP subtypes (S1-S6) are visualised by the time when 25%, 50%, and 75% of patients evaluated for a particular symptom developed it. F, frontal presentation; Misc, miscellaneous presentations; OM, ocular motor dysfunction; P, parkinsonism; PF, postural and frontal dysfunction; PI, postural instability; PSP, progressive supranuclear palsy; RS, Richardson’s syndrome; SL, speech/language disorders

### Neuropathologic characteristics of early PSP subtypes

The total tau score (range: 0–12) revealed significant regional variations across the early PSP subtypes (Table 2). PSP-PF exhibited higher total tau burdens in both cortical and subcortical regions, including the superior frontal cortex (7 [5, 9], P < 0.01), thalamic fasciculus (6 [5, 6], P < 0.01), red nucleus (10 [8, 11], P = 0.01), midbrain tectum (11 [9, 11], P = 0.03), pontine base (6 [5, 7], P = 0.01), and cerebellar white matter (4 [3, 6], P < 0.01). PSP-SL showed high total tau scores in the superior frontal cortex (9 [6, 10], P < 0.01) and in the temporal cortex (3 [2, 6], P = 0.01). Frequencies of argyrophilic grains showed no significant differences between subtypes. TDP-43 pathology in the amygdala was more frequently observed in PSP-F than in PSP-PF (19.2% vs. 3.2%, P < 0.05) and tended to be frequent in PSP-SL (17.6%) as well (Table 1).

**Table 2.** Total tau score between subtypes.

|  | Overall | Subtypes |  |  |  |  |  |  |  | *P*<br>value |
| --- | --- | --- | --- | --- | --- | --- | --- | --- | --- | --- |
|  |  | PSP-PF | PSP-RS | PSP-PI | PSP-P | PSP-SL | PSP-F | PSP-OM | Misc |  |
| *n* | 588 | 188 | 68 | 204 | 36 | 29 | 31 | 13 | 19 |  |
| Superior frontal cortex | 7 [5, 8] (362) | 7 [5, 9]<br>(127)<sup>OM,P,PI</sup> | 6 [5, 8] (37) | 6 [4, 8] (118)<sup>PF</sup> | 5 [3, 7] (26)<sup>PF,SL</sup> | 9 [6, 10] (19)<sup>OM,P</sup> | 7 [6, 8] (15) | 4 [2, 6] (9)<sup>PF,SL</sup> | 7 [5, 10] (11) | **<0.01** |
| Motor cortex | 9 [7, 11] (559) | 10 [7, 11] (180) | 9 [8, 10] (65) | 9 [6, 11] (194) | 8 [5, 10] (36) | 9 [8, 11] (26) | 9 [6, 10] (27) | 8 [3, 10] (13) | 10 [8, 11] (18) | **0.02** |
| Temporal cortex | 2 [1, 3] (561) | 2 [1, 3] (180) | 2 [1, 3] (65)<sup>SL</sup> | 2 [1, 3] (195) | 2 [1, 3] (36) | 3 [2, 6] (27)<sup>OM,PF</sup> | 2 [1, 3] (27) | 1 [1, 2] (13)<sup>SL</sup> | 3 [1, 4] (18) | **<0.01** |
| Caudate/putamen | 8 [6, 9] (562) | 8 [6, 9] (180) | 8 [6, 9] (65) | 7 [6, 9] (196) | 6 [6, 8] (36) | 8 [7, 9] (27) | 8 [7, 8] (27) | 6 [5, 9] (13) | 7 [6, 8] (18) | 0.11 |
| Globus pallidus | 7 [6, 9] (559) | 8 [6, 9] (180) | 7 [6, 9] (65) | 7 [6, 9] (194) | 7 [5, 9] (36) | 7 [5, 9] (27) | 8 [6, 9] (26) | 8 [7, 9] (13) | 7 [6, 10] (18) | 0.31 |
| Basal nucleus | 7 [6, 8] (560) | 7 [6, 8] (180) | 7 [5, 7] (65) | 7 [5, 7] (194) | 6 [6, 8] (36) | 7 [5, 8] (27) | 7 [5, 8] (27) | 6 [4, 7] (13) | 7 [5, 9] (18) | 0.43 |
| Hypothalamus | 5 [4, 6] (551) | 5 [4, 6] (176) | 5 [4, 6] (65) | 5 [4, 6] (193) | 4 [4, 6] (35) | 6 [3, 6] (25) | 4 [4, 6] (27) | 4 [4, 5] (12) | 4 [3, 5] (18) | 0.57 |
| Ventral thalamus | 9 [8, 10] (560) | 9 [8, 10] (180) | 9 [8, 10] (65) | 9 [8, 10] (195) | 9 [6, 10] (36) | 9 [8, 10] (27) | 10 [8, 10] (26) | 9 [7, 10] (13) | 9 [8, 10] (18) | 0.19 |
| Subthalamic nucleus | 9 [8, 11] (562) | 10 [8, 11] (180) | 9 [8, 11] (65) | 9 [8, 10] (196) | 8 [8, 10] (36) | 8 [6, 10] (27) | 10 [8, 11] (27) | 9 [7, 10] (13) | 10 [8, 11] (18) | 0.03 |
| Thalamic fasciculus | 5 [4, 6] (560) | 6 [5, 6] (179)<sup>OM,P</sup> | 5 [5, 6] (65) | 5 [4, 6] (196) | 5 [4, 6] (35)<sup>PF</sup> | 6 [4, 6] (27) | 5 [4, 6] (27) | 4 [2, 5] (13)<sup>PF</sup> | 6 [4, 6] (18) | **<0.01** |
| Red nucleus | 9 [7, 10] (560) | 10 [8, 11] (180)<sup>P</sup> | 9 [7, 10] (65) | 9 [7, 10] (194) | 8 [4, 9] (36)<sup>PF</sup> | 9 [6, 11] (27) | 9 [8, 10] (27) | 9 [5, 11] (13) | 9 [7, 10] (18) | **0.01** |
| Substantia nigra | 7 [6, 8] (560) | 7 [6, 9] (179) | 7 [6, 9] (65) | 7 [6, 8] (195) | 6 [5, 8] (36) | 6 [6, 8] (27) | 8 [7, 8] (27) | 6 [6, 7] (13) | 8 [6, 9] (18) | **0.01** |
| Oculomotor complex | 6 [4, 7] (500) | 6 [5, 7] (157) | 5 [4, 7] (57) | 6 [4, 7] (173) | 5 [4, 6] (34) | 5 [4, 6] (26) | 6 [5, 7] (25) | 5 [3, 6] (11) | 5 [3, 6] (17) | 0.07 |
| Midbrain tectum | 10 [9, 11] (557) | 11 [9, 11]<br>(180)<sup>OM</sup> | 10 [9, 11] (64) | 10 [9, 11] (194) | 10 [8, 11] (35) | 10 [9, 11] (26) | 11 [10, 12] (27) | 8 [8, 9] (13)<sup>PF</sup> | 10 [9, 11] (18) | **0.03** |
| Locus coeruleus | 6 [5, 6] (532) | 6 [5, 6] (172) | 6 [5, 6] (62) | 6 [5, 6] (185) | 6 [5, 6] (32) | 5 [5, 6] (27) | 6 [5, 6] (25) | 6 [5, 6] (12) | 6 [5, 6] (17) | 0.19 |
| Pontine tegmentum | 8 [7, 9] (555) | 8 [7, 9] (179) | 7 [7, 8] (64) | 7 [7, 8] (195) | 7 [6, 8] (34) | 8 [6, 8] (26) | 8 [7, 9] (26) | 7 [5, 8] (13) | 8 [7, 9] (18) | 0.09 |
| Pontine base | 6 [5, 7] (558) | 6 [5, 7] (180)<sup>P</sup> | 6 [6, 7] (65)<sup>P</sup> | 6 [5, 7] (195) | 5 [3, 6] (34)<sup>PF,RS</sup> | 7 [6, 8] (27) | 6 [5, 7] (26) | 6 [2, 7] (13) | 6 [5, 7] (18) | **0.01** |
| Medullary tegmentum | 7 [7, 8] (533) | 7 [7, 8] (173) | 7 [7, 8] (63) | 7 [7, 8] (184) | 7 [6, 8] (33) | 7 [5, 8] (27) | 8 [7, 8] (25) | 7 [6, 7] (11) | 8 [7, 8] (17) | 0.1 |
| Inferior olive | 6 [5, 7] (522) | 6 [5, 7] (168) | 6 [5, 7] (62) | 6 [5, 7] (183) | 6 [5, 7] (31) | 6 [4, 7] (26) | 6 [6, 8] (25) | 5 [4, 6] (11) | 6 [5, 7] (16) | 0.38 |
| Dentate nucleus | 5 [4, 6] (556) | 5 [4, 6] (179) | 5 [4, 6] (65) | 5 [4, 6] (194) | 5 [4, 6] (35) | 5 [3, 6] (26) | 5 [4, 7] (26) | 5 [3, 6] (13) | 5 [4, 7] (18) | 0.92 |
| Cerebellar white matter | 4 [3, 5] (553) | 4 [3, 6] (179)<sup>OM,P</sup> | 4 [3, 5] (65) | 4 [3, 5] (192) | 3 [2, 4] (34)<sup>PF</sup> | 4 [2, 5] (26) | 4 [3, 5] (26) | 2 [1, 3] (13)<sup>PF</sup> | 4 [3, 5] (18) | **<0.01** |

The total tau score (0-12) is defined as the sum of four semi-quantitative tau pathology scores (0-3 each): neurofibrillary tangles/pretangles, coiled bodies, tufted astrocytes and neuropil threads. Values are presented as median [25th and 75th percentiles] (number of cases evaluated). Overall statistical differences are evaluated using the Kruskal-Wallis test, and the corresponding *P*-values are presented, followed by the pairwise Steel-Dwass test for variables with significance. *P* value < 0.05 is considered statistically significant. Superscripts PF, RS, PI, P, SL, F, OM, and Mi indicate *P* < 0.05 vs. subtype PSP-PF, PSP-RS, PSP-PI, PSP-P, PSP-SL, PSP-F, PSP-OM, and Misc, respectively. F, frontal presentation; Misc, miscellaneous presentations; OM, ocular motor dysfunction; P, parkinsonism; PF, postural and frontal dysfunction; PI, postural instability; PSP, progressive supranuclear palsy; RS, Richardson's syndrome; SL, speech/language disorder

## Discussion

In the present study, a fully automated pipeline integrating fine-tuned ChatGPT at three steps—indexing, symptom identification, and onset estimation—screened 53,527 pages of medical records from 1,344 autopsy-confirmed patients with PSP and abstracted 195 clinical presentations with onset dates. Using early presentations from 588 cases with qualified records, UMAP clustering identified six early subtypes. A simplified decision tree model was developed to efficiently incorporate these subtypes. Using five clinical features, the decision tree model successfully classified early PSP subtypes in a mutually exclusive manner, which corresponded to subsequent clinical progression, prognoses, and post-mortem pathology. The clinical value of the subtyping was supported by a statistically meaningful improvement in prognostic prediction; incorporating the subtyping information increased the C-index from 0.554 to 0.654 (Δ = 0.100). Although 3% (19/588) of patients did not exhibit any of the five core presentations, this proportion is improved from the 13% of patients who remained unclassified with the Respondek criteria.[14]

Our models identified a distinct presentation that has not been described in previous literature, characterized by early postural and frontal dysfunction, which we termed PSP-PF. This subtype presents with early cortical and subcortical symptoms, rapid progression, the shortest disease duration, and high tau burden in both the cortical and subcortical regions on postmortem analysis. Respondek et al. reported that approximately two-thirds of patients with PSP-RS had frontal presentations three years post-onset.[14] In the present cohort, the majority of PSP-PF patients who were evaluated for OM within three years of onset had it. (83.0%, 122/147, Supplementary Table 6) These patients would have been classified as PSP-RS under the MDS criteria and MAX rule due to the presence of both OM and PI. [10, 41, 42] However, the MDS criteria and MAX rule have come under scrutiny for potentially underestimating the phenotypic diversity of PSP due to prioritizing PSP-RS over other phenotypes, including the PSP-F subtype.[11–13] The recognition of PSP-PF as an aggressive variant of PSP in this simple decision tree model may address these concerns and help clinicians differentiate subtypes and estimate prognoses at the bedside.

PSP-PI was defined by presence of PI and absence of OM or frontal presentation within three years, consisting of the largest group of early PSP subtypes. Most patients who presented with PSP-PI early in the disease course were reported to subsequently develop OM and therefore transition to PSP-RS,[13, 14] which aligns with current observations (Fig. 3). The PSP-PI subtype had a longer disease duration than PSP-RS, without significant differences in lifelong symptoms or postmortem tau evaluation. This supports the idea that PSP-PI and PSP-RS may share common characteristics but have different speeds of progression, and that most patients presenting with PSP-PI could be in either a premature or underestimated state of PSP-RS.

PSP-P in the current study was defined as the presence of any parkinsonian features (bradykinesia, rigidity, and tremor) and the absence of frontal presentation, PI, or SL within three years of disease onset. Levodopa responsiveness in PSP-P could not be determined due to the small number of records containing this information (3/36 were assessed, shown in Supplementary Table 6). PSP-P had the longest disease duration among the subtypes, with 10 years [8, 10]. Pathological findings included a low tau burden in the superior frontal cortex. These findings align with previous reports on PSP-P. [7, 9, 13, 40, 43]

Patients with PSP-SL initially developed nonfluent/agrammatic primary progressive aphasia and/or progressive apraxia of speech.[9] As years passed, they developed other cortical symptoms and parkinsonian features, resulting in a relatively longer disease duration of eight years [7, 10]. Postmortem analysis revealed high tau burden in the superior frontal and temporal cortexes. These clinicopathologic features are consistent with previous reports.[13, 43–48] As many individuals in the current cohort with PSP-SL did not develop additional symptoms until approximately four years after onset, patients with isolated speech and language dysfunction should be carefully monitored for the possibility of PSP.[49]

Patients with PSP-F initially presented with at least one behavioral or dysexecutive feature (referred to here as frontal presentation). Approximately four-to-five years later, half of patients developed parkinsonian features and upper or lower motor neuron signs. PSP-F had the second-longest disease duration of nine years [7, 10] after PSP-P. Previous studies have indicated that PSP-F had a relatively shorter disease duration[14] and a tendency to be unclassified by the MDS-PSP criteria in the early stage.[13] These differences could originate from stricter definitions of frontal presentations; the Respondek and MDS-PSP criteria require a minimum of two and three frontal symptoms, respectively. Our study adopted one as the threshold for the number of the frontal presentations because early behavioral changes were often summarised in the records by the family or clinicians with a single description such as “personality change” or “apathy.” Additionally, when a higher threshold was tested, this generated many missing values, as indicated with “Frontal presentation (≥3)” (Supplementary Table 6). By implementing different criteria, the present study broadly captured an earlier cluster of PSP-F in which frontal presentation preceded other symptoms by five years. The higher prevalence of TDP-43 pathology in the amygdala in PSP-F compared with PSP-PF (19.2% vs. 3.2%, P < 0.05; Table 1) suggests a potential contribution of TDP-43 pathology to the clinical heterogeneity of PSP. To elucidate the underlying mechanisms, we acknowledge the importance of further investigation, including TDP-43 immunohistochemistry in the cerebral cortex.

In the current study, PSP-OM was defined as patients presenting with OM without frontal presentation, PI, or SL within three years of disease onset, constituting the smallest subgroup during the early stage. Patients classified as PSP-OM in this cohort subsequently developed cortical signs, parkinsonian signs, and PI, leading to a moderate disease duration (Figs. 2d and 4). This finding is consistent with previous literature that reported patients with PSP-OM subsequently develop PI.[9, 14] However, due to the small number of cases, it remains uncertain whether PSP-OM belongs to the spectrum of PSP-RS.

The current study did not identify PSP-CBS as an independent subtype. Among 79 patients who had cortical signs, 50 patients were assigned to PSP-PF, nine to PSP-RS, seven to PSP-PI, and six to PSP-SL (P = 0.10, supplementary Table 6), suggesting that most PSP patients with cortical signs at an early stage also exhibit other presentations, such as OM, PI, or SL. This fits the earlier studies on PSP-CBS, in which PSP-CBS was defined without excluding PI or OM, and many PSP-CBS patients were reported to have or subsequently developed PI or OM.[14, 50, 51]

The current pipeline successfully abstracted presence of symptoms paired with onset information, by dividing the task into three simple steps and fine-tuning the LLM at each step with a large set of excerpts with human annotation. This approach has not been reported in previous applications of LLM to clinical data collection.[17–21] Our pipeline has several strengths. Its reliability was validated with human annotations at each step. The medical records used in the study comprised different document formats from different institutions, and the pipeline can therefore be easily adapted for use in other studies. The abstraction results were interpretable because they are created by a rule-based algorithm integrating three simple tasks. This method can be applied to data collection for a variety of purposes, such as characterizing the progression of other neurodegenerative diseases, or establishing diagnostic criteria at a specific timepoint.

This retrospective study, which primarily focused on analysing a large clinicopathologic dataset, has several limitations. Because our pipeline was designed to abstract symptom presence, it could not assess objective values such as neuropsychological test scores and L-dopa dosage; these should be incorporated in future studies. As the majority of subjects in this study were Caucasian, future validation with cases from diverse demographics is needed, particularly in PSP-C, which is suspected to differ in frequency by race.[26, 27] Documentation practices in clinical notes were not standardized and may have varied over the long retrospective period (1998–2022), resulting in some parameters being unavailable. Unavailable data were excluded from each analysis, except in the UMAP analysis, where those were replaced with zeros due to methodological constraints. As the handling of missing data may affect the results, the total number of cases and the number of cases evaluated are listed together in the tables to aid interpretation. Still, we believe the reliability of the clinical data is supported by the high proportion of specialist evaluations (99.1%) and by the requirement for symptom descriptions at least at two time points—within three years of disease onset and after three years. For symptoms or signs without a documented time of onset, the first available evaluation date was considered the onset date. While this approach is reasonable given that many parameters are based on findings from neurological examinations, it may not accurately capture precise onset information. This is a general limitation of retrospective studies and is not specific to the automated abstraction of clinical features.

### Conclusions

Using a large-scale postmortem dataset, this data-driven study identified a novel and aggressive variant of PSP, characterized by early postural and frontal dysfunction. A simple decision tree model would help clinicians diagnose diverse PSP phenotypes, estimate disease progression, and facilitate early patient recruitment for clinical trials.

## Acknowledgements

The authors would like to thank all patients and their families for their agreement to brain donation, and Rachel R. LaPaille-Harwood and Audrey J. Strongosky (Mayo Clinic, Jacksonville, FL) for assistance with brain banking, and to Monica Castanedes-Casey (Mayo Clinic, Jacksonville) for histologic support.

## Author contributions

DO conceived the study concept and design, annotated the datasets, analysed the results, and drafted the manuscript. HS, NBG, and SFR interpreted and discussed the results. ARM acquired and annotated the datasets. RJU, IL, KAJ, and ZKW acquired the clinical data and reviewed the manuscript. DWD reviewed the manuscript and supervised the study. All authors commented on and approved the final manuscript.

## Data Availability

The data and codes used in the current study are available from the corresponding author upon reasonable request.

