## Supplemental Information for "Early Subtypes and Progressions of Progressive Supranuclear Palsy: A Data-Driven Brain Bank Study"

Daisuke Ono, MD, PhD^1,2,3^, Hiroaki Sekiya, MD, PhD^1^, Nikhil B. Ghayal, BS^1^, Alexia R. Maier, BS^1^, Shanu F. Roemer, MD, PhD^1^, Ryan J. Uitti, MD^4^, Irene Litvan, MD^5^, Keith A. Josephs, MD, MST, MSc^6^, Zbigniew K. Wszolek, MD^3^, and Dennis W. Dickson, MD^1^

^1^Department of Neuroscience, Mayo Clinic, Jacksonville, Florida, USA

^2^Department of Neurology and Neurological Science, Institute of Science Tokyo, Tokyo, Japan

^3^School of Statistical Thinking, The Institute of Statistical Mathematics, Tokyo, Japan

^4^Department of Neurology, Mayo Clinic, Jacksonville, Florida, USA

^5^Department of Neurosciences, University of California, San Diego, California, USA.

^6^Department of Neurology, Mayo Clinic, Rochester, Minnesota, USA.

Correspondence to: Daisuke Ono and Dennis W. Dickson

Department of Neuroscience, Mayo Clinic, 4500 San Pablo Road, Jacksonville, FL 32224, USA

 (DO) and (DWD)

**Supplementary Methods**

**Neuropathologic procedures**

Brain autopsies were performed with the consent of legal next of kin or individuals with legal authority to grant permission for autopsy. Formalin-fixed brains were sampled using a standardised method by a board-certificated neuropathologist (DWD).[1] Paraffin-embedded 5-μm thick sections mounted on glass slides were stained with hematoxylin & eosin and thioflavin S fluorescence microscopy. Immunohistochemistry was performed for phosphorylated-tau (CP13; mouse monoclonal, 1:1000; from the late Dr. Peter Davies, Feinstein Institute for Medical Research, NY) and phosphorylated TDP-43 (p409/410; mouse monoclonal, 1:5,000; CosmoBio USA, Carlsbad, CA or MC2085, rabbit polyclonal, 1:3,000; from Dr Leonard Petrucelli, Mayo Clinic, Jacksonville, FL) using an IHC Autostainer (Thermo Fisher Scientific, Waltham, MA, USA) with Dako EnVisionTM + reagents (Agilent Technologies, Santa Clara, CA) and 3, 3-diaminobenzidine (DAB) as the chromogen with counterstaining with haematoxylin.

**Fine-tuning ChatGPT**

Patients' records in the Mayo Clinic Brain Bank obtained by their next-of-kin or other person with close personal ties to the patient. For Mayo Clinic patients, clinical information was abstracted form the electronic medical record or from PDF files derived from the electronic medical record. Text data were extracted using an optical character recognition library, python-tesseract 0.3.10, and then de-identified using a natural language processing library, spaCy 3.7.2. For each task, excerpts from the medical records were annotated by ARM and DO and randomly split into training and test datasets. Fine-tuning of gpt-4o-mini-2024-07-18 from the ChatGPT API openai 1.51.2 was performed with hyperparameters of n_epochs:3, batch_size:1 and learning_rate_multiplier:2. The prompts used in the fine-tuning were detailed in Supplementary Table 1. The model performance was evaluated using the accuracy of the hold-out test dataset at each step.

1) **Indexing patients’ documents**

The patients’ records in this study contained different types of documents in various formats from different sources, and included family questionnaires, test results, consent forms, as well as medical records. To identify the type of document and date of evaluation for each page, ChatGPT was asked to create an index of each document.

To generate an input dataset for fine-tuning, 632 documents from 581 patients were randomly selected from the Mayo Clinic Brain Bank. The type of document and date of evaluation were manually determined. The annotated dataset was randomly divided into 570 training and 62 test documents. The first and last five lines, in addition to those including the date information in the remaining part, were excerpted from each page and used for fine-tuning.

2) **Determining the presence of symptoms**

To capture the diverse clinical presentations of PSP, 195 medical histories, symptoms, signs, and their search terms were selected, with reference to previous literature and the MDS-PSP criteria (Supplementary Table 2). [10–13] The clinicians' notes and test results, identified in the previous indexing step, were screened for the search terms, and three lines containing any of the terms were then excerpted. From the medical records of 789 donors, 4,245 excerpts containing a search term were randomly selected. Reviewers assessed whether a patient had, did not have, or could not be determined to have the clinical presentation based on an excerpt. The annotated dataset was randomly divided into 4,145 training and 100 test excerpts, which were used for fine-tuning and performance evaluation, respectively.

3) **Identifying the onset of symptoms**

Using the algorithms above, indices and information on symptom presence were obtained from 757 randomly selected donors, from which 4,029 excerpts with evaluation of a symptom were randomly selected for a fine-tuning dataset. Reviewers assessed these excerpts using four formats for symptom onset: 1) date, 2) age, 3) month (and day) in year of recording, and 4) years since date of recording. The annotated dataset was randomly divided into 3,554 training and 475 test excerpts, which were used for fine-tuning and performance evaluation, respectively. The date of symptom onset was calculated using the prediction of four onset formats, date of birth, and the date of evaluation from the indices generated in the previous step. Accuracy for the year and month was calculated, ignoring the consistency of the day, given the chronic nature of the neurodegenerative disease.

**Clinical record abstraction**

An automated pipeline for the abstraction of structured clinical records was developed (Supplementary Fig. 1). GPT-4o was fine-tuned with human-annotated datasets for three tasks: indexing patient documents, identifying symptom presence, and determining symptom onset. The models achieved accuracies of 0.90 for document date abstraction, 0.95 for document type, 0.89 for symptom presence, and 0.94 for symptom onset, which confirmed feasibility for further analysis.

**Summarizing structural clinical data**

Clinical presentations obtained by the fine-tuned GPT were summarised for each patient according to the following workflow: 1) the label “undetermined” was ignored, 2) if a patient had at least one "present" label for a symptom during an assessment period (within 3 years of disease onset or lifelong), then the patient was considered to have the symptom, 3) if all the label(s) showed “absent”, then the patient was considered not to have the symptom, and 4) if a patient did not have any label, the value was marked as “not available.” The same presentations from different expressions were grouped, and syndromes important for PSP were defined by modifying from previous literature, [10–13] as detailed in Supplementary Table 3.

The onset of each symptom/sign was determined as follows: 1) the date of first available evaluation was considered as the date of onset if there was no description of onset. 2) if the symptom or sign was present, the earliest date recorded was adopted. Symptom duration was calculated by subtracting the symptom onset from disease onset. To be consistent with the MDS-PSP criteria and its validation studies, we considered symptoms and signs within three years of disease onset as early presentations.[13–15]

**Supplementary Figure**

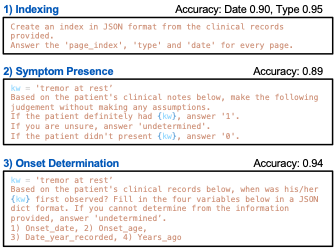

**Supplementary Fig. 1 Abstraction of structured clinical records with fine-tuned ChatGPT**

The pipeline for abstracting structured clinical records involves three steps: 1) indexing patient documents, where the type and date of each document are extracted for each page, 2) determining the presence of symptoms, with ‘tremor at rest’ as an example, and 3) identifying the onset of symptoms, where the onset date for each symptom is determined. GPT-4o is fine-tuned using excerpts from the records, with human-labelled annotations serving as the gold standard. The complete prompts are provided in Supplementary Table 1

**Supplementary Tables**

**Supplementary Table 1 Prompts used for the ChatGPT-integrated medical record abstraction**

1) Indexing

Create an index in JSON format from the clinical records provided.

The documents consist of patient questionnaires, content forms, clinical notes, test results, etc. Each document is filed in an orderly manner.

Their headers and footers are de-identified and abstracted for your review.

Answer the ‘page_index’, ‘type’ and ‘date’ for every page.

The page number should match the “PAGE_INDEX ~ OF ENTIRE DOCUMENT” written on the first line of each page.

You need to select “date” from the “dates” list, which is abstracted and provided beforehand to avoid hallucination.

We need “Date” means date of service (DOS), date of encounter, date of clinician signature, or date of examination, not dates when documents were printed or sent.

2) Symptom Presence

You only need to judge based on the following patient’s clinical records. Do not hallucinate.

If the patient definitely had {kw}, answer ‘1’.

If the patient didn’t present {kw}, answer ‘0’.

If you are unsure or it mentions only a risk of {kw}, answer ‘undetermined’.

3) Onset Determination

In case {kw} was present.

Based on the patient’s clinical records below, when was his/her {kw} first observed?

Fill in the three variables below in a JSON dict format. If you cannot determine from the information provided, answer ‘undetermined’.

1) Onset_date: Answer the date in MM/DD/YYYY when {kw} was observed. Do not answer the printed date.

2) Onset_age: Answer how old the patient was when {kw} was observed.

3) Date_year_recorded: If {kw} was observed in the year recorded, answer the observed month and day in MM/DD/yyyy.

4) Years_ago: Answer how many years before the recording date when {kw} was observed.

In case {kw} was absent.

Based on the patient’s clinical records below, when was the absence of {kw} observed?

Fill in the three variables below in a JSON dict format. If you cannot determine from the information provided, answer ‘undetermined’.

1) Onset_date: Answer the date in MM/DD/YYYY when the absence of {kw} was observed. Do not answer the printed date.

2) Years_ago: Answer how many years before the recording date when the absence of {kw} was observed.

3) Date_year_recorded: If the absence of {kw} was observed in the year recorded, answer the observed month and day observed in MM/DD/yyyy.

4) Onset_age: Answer how old the patient was when the absence of {kw} was observed.

**Supplementary Table 2 Search and exclusion terms for the automated collection of 195 symptoms and signs**

| Symptoms / signs | Search terms | Exclusion terms |
| --- | --- | --- |
| hypomimia | hypomimia, amimia |  |
| masked face | masked face |  |
| bradykinesia | bradykine |  |
| slow movement | slow mov |  |
| decreased arm swing | arm swing |  |
| micrographia | micrograph |  |
| small handwriting | hand writ |  |
| akinesia | akine |  |
| no spontaneous movements | spontaneous mov |  |
| gait freezing | gait\|walk freezing |  |
| glued gait | glued |  |
| magnetic gait | magnetic gait\|walk |  |
| rigidity | rigid |  |
| axial rigidity | axi rigid |  |
| nuchal (neck) rigidity | nuchal rigid, neck rigid |  |
| appendicular (limb) rigidity | appendicular rigid, limb rigidity |  |
| cogwheel rigidity | cogwheel |  |
| stiffness | stiffness |  |
| increased muscle tone | muscle tone |  |
| the fact patient fell, not risk of fall | fall, fell |  |
| early falls | early falls |  |
| multiple/frequent falls | multi\|frequent\|often fall\|fell |  |
| falling forwards | fall\|fell forward |  |
| positive pull test | pull test |  |
| falling backwards | fall\|fell backward |  |
| imbalance | imbalance |  |
| shuffling gait | shuffl gait |  |
| festinating gait | festinat gait |  |
| turning en bloc | en bloc |  |
| stooped posture | stoop, bent, bend |  |
| pisa sign | pisa |  |
| camptocormia | camptocormia |  |
| scoliosis | scolio |  |
| memory loss//deficit | memory, forget, recall | clinic |
| memory loss/deficit benefited by cues | cueing\|cues |  |
| amnesia | amnes |  |
| word finding difficulty | word find |  |
| disorientation | orient |  |
| lost date | lost date |  |
| lost name | lost name |  |
| lost place | lost place |  |
| impaired serial sevens | serial sevens\|7 |  |
| abnormal clock test | clock test |  |
| impaired spelling WORLD backwards | spell\|WORLD backwards |  |
| wander | wander |  |
| gets lost | get\|got lost |  |
| prosopagnosia | prosopagnosia |  |
| unable to answer famous faces | famous\|president face |  |
| fluctuation of neuropsychiatric symptoms | fluctuat |  |
| sundowning | sundown |  |
| hallucinations | hallucinat |  |
| visual hallucinations | hallucinat |  |
| paranoid delusions | paranoi |  |
| apathy | apathy |  |
| social withdrawal | social isolation, withdraw, reclusive |  |
| abulia | abulia |  |
| bradyphrenia | bradyphrenia, thinking |  |
| change in personality | personality |  |
| disinhibition | disinhibit |  |
| impulsivity | impuls |  |
| obsession | obsess |  |
| compulsivity | compulsiv |  |
| hyperorality | hyperoral |  |
| abnormally increased appetite | appetite |  |
| hyperphagia | hyperphagia |  |
| perseveration | persever |  |
| echolalia | echolalia |  |
| palilalia | palilalia |  |
| groping | groping |  |
| speaking out of turn | interject, interrupt, speak\|spoke out of turn |  |
| stereotyped behavior | stereotyp |  |
| joking/jovial behavior | joke\|joking, clown, jovial |  |
| kleptomania/stealing | kleptomania, steal |  |
| cussing behavior | cussing, los filter |  |
| hypersexuality | libido, sexual |  |
| caught in a scam/fraud | scam, fraud |  |
| gambling | gambling |  |
| addiction/abuse | addict, abuse |  |
| Baker acted | Baker act |  |
| suicidal ideation | suicid |  |
| criminal behavior | jail, convict, criminal\|crime | discriminat |
| hospitalized in psychiatric ward/asylum | psychiatric ward\|asylum\|hospital |  |
| positive grasping reflex | grasp |  |
| positive snout reflex | snout |  |
| positive sucking reflex | sucking |  |
| positive palmomental reflex | palmomental |  |
| frontal release sign | frontal release |  |
| gegenhalten | gegenhalten |  |
| paratonia | paratoni |  |
| abnormal Luria test | Luria test\|sequence |  |
| abnormal fist-edge-palm test | fist edge palm test\|sequence |  |
| applause sign | applause, clap |  |
| abnormal go-no-go test | go no test |  |
| stopped driving | driv stop\|license |  |
| car accident | car accident |  |
| metamorphosis | metamorphosis |  |
| loss of appetite | appetite |  |
| cries a lot | crie, cry |  |
| aphasia/primary progressive aphasia (PPA) | aphasia, PPA |  |
| agrammatic aphasia | agrammatic aphasia |  |
| reduced fluency | fluent\|fluenc |  |
| apraxia of speech | apraxia of speech |  |
| abnormal articulation/pronunciation | articulat, pronunciat |  |
| jumbled speech | jumbled |  |
| spasmodic speech/dysphonia | spasmodic |  |
| mutism | mute\|mutism |  |
| alien hand or limb | alien |  |
| orobuccal apraxia | orobuccal |  |
| apraxia | apraxia | apraxia of speech |
| ideomotor apraxia | ideomotor apraxia |  |
| cannot pantomime | pantomime |  |
| limb apraxia | limb apraxia |  |
| unable to stick out tongue | stick tongue |  |
| unable to lick lips | lick lip |  |
| unable to whistle | whistl |  |
| simultanagnosia | simultanagnosia |  |
| cortical sensory deficit | cortical sensory\|sense |  |
| REM sleep behavior disorder (RBD) | RBD, REM, dream enact |  |
| hyposmia | hyposmia |  |
| loss of smell | smell, olfactory |  |
| restricted eye movement/gaze palsy | EOM, extraocular, gaze |  |
| vertical hypometric saccades | hypometri |  |
| downgaze palsy | downgaze |  |
| slow saccades | saccad |  |
| eyelid opening apraxia | eyelid opening |  |
| blepharospasm | blepharospasm |  |
| photophobia | photophobia |  |
| ataxia | atax | ataxia clinic |
| hypermetric saccades | hypermetri saccad |  |
| saccadic hypometria | hypometri saccad |  |
| overshoot | overshoot |  |
| scanned speech | scanned |  |
| dysmetria | dysmetri |  |
| hypermetria | hypermetri |  |
| abnormal finger to nose | finger nose, FNF\|FN test |  |
| abnormal heel to shin | heel shin, HS test |  |
| positive or abnormal Romberg sign | Romberg |  |
| square-wave jerks | square jerk |  |
| nystagmus | nystagmus |  |
| dysdiadochokinesis | diadocho |  |
| unable to walk a line | walk line |  |
| unable to perform tandem gait | tandem gait |  |
| involuntary movement | writhing, twitch, jerk | square\|ankle |
| chorea/writhing | chore |  |
| athetosis | atheto |  |
| dyskinesia | dyskinesia |  |
| hemifacial spasm | hemifacial spasm |  |
| myokymia | myokymia |  |
| shaking | shak |  |
| tremor | tremor |  |
| tremor at rest | rest tremor |  |
| postural tremor | postur tremor |  |
| kinetic tremor | kinet tremor |  |
| essential tremor (ET) | essential tremor, ET | et al |
| jaw/chin tremor | jaw\|chin tremor |  |
| myoclonus | myoclon |  |
| cortical myoclonus | cortical myoclon |  |
| limb myoclonus | limb myoclon |  |
| facial myoclonus | fac myoclon |  |
| finger myoclonus | finger myoclon |  |
| polyminimyoclonus | polyminimyoclonus |  |
| dystonia | dystoni |  |
| antecollis | antecollis |  |
| retrocollis | retrocollis |  |
| clenched fist | clench fist |  |
| hand dystonia | dyston hand |  |
| foot dystonia | dyston foot |  |
| upper motor neuron (UMN) signs | upper motor neuron, UMN |  |
| pseudobulbar palsy | pseudobulbar |  |
| inappropriate/pathological laughter | laugh |  |
| inappropriate/pathological crying | crie, cry |  |
| hyperreflexia | hyperreflex |  |
| positive Hoffmann sign/reflex | Hoffmann |  |
| positive/extensor/upgoing (not absent/flexor/downgoing) Babinski sign/reflex | Babinski |  |
| positive/extensor/upgoing (not absent/flexor/downgoing) plantar reflex | plantar |  |
| spasticity | spastic |  |
| lower motor neuron (LMN) signs | upper motor neuron, LMN |  |
| fasciculations | fasciculat |  |
| tongue atrophy | tongue atroph\|muscle |  |
| dysarthria | dysarthria |  |
| dysphasia | dysphasia |  |
| swallowing difficulty | swallow |  |
| aspiration | aspirat |  |
| PEG/feeding tube insertion | PEG, feeding tube |  |
| stridor | stridor |  |
| lightheaded/dizziness | lighthead, dizz |  |
| orthostatic hypotension | orthostatic |  |
| urinary dysfunction | urinar |  |
| urinary incontinence | incontinence |  |
| urinary catheter | catheter |  |
| urinary tract infection (UTI) | urinary infection, UTI |  |
| erectile dysfunction (ED) | erectile |  |
| abnormal sweat | sweat |  |
| cold discolored hands/feet | discolored\|cold hand\|foot\|feet |  |
| improvement with l-dopa, levodopa or sinemet | l l-dopa\|levo\|sinemet benefit\|respon\|improv |  |

**Supplementary Table 3 Definition of the syndromes**

| Syndrome | Criteria |
| --- | --- |
|  | At least 1 of the following items |
| Ocular motor dysfunction | restricted eye movement/gaze palsy, vertical hypometric saccades, downgaze palsy, slow saccades, eyelid opening apraxia, blepharospasm |
| O1-2 (MDS 2017) | restricted eye movement/gaze palsy, vertical hypometric saccades, downgaze palsy, slow saccades |
| Postural instability | early falls, multiple/frequent falls, falling backwards |
| P1-2 (MDS 2017) | early falls, multiple/frequent falls, falling backwards, positive pull test |
| Akinesia | akinesia, no spontaneous movements, gait freezing, glued gait, magnetic gait |
| Bradykinesia | bradykinesia, slow movement, hypomimia, masked face |
| Rigidity | rigidity, axial rigidity, nuchal (neck) rigidity, appendicular (limb) rigidity, cogwheel rigidity, increased muscle tone |
| Rest tremor | tremor at rest, jaw/chin tremor |
| Any parkinsonian gait | shuffling gait, festinating gait, turning en bloc |
| Frontal presentation | apathy, social withdrawal, abulia, bradyphrenia, change in personality, disinhibition, impulsivity, obsession, compulsivity, hyperorality, abnormally increased appetite, hyperphagia, perseveration, echolalia, palilalia, groping, speaking out of turn, stereotyped behavior, joking/jovial behavior, kleptomania/stealing, cussing behavior, hypersexuality, caught in a scam/fraud, gambling, addiction/abuse, Baker acted, suicidal ideation, criminal behavior, hospitalized in psychiatric ward/asylum, positive grasping reflex, positive snout reflex, positive sucking reflex, positive palmomental reflex, frontal release sign, gegenhalten, paratonia, abnormal Luria test, abnormal fist-edge-palm test, applause sign, abnormal go-no-go test |
| Any abnormal posture | stooped posture, pisa sign, camptocormia, scoliosis |
| Cortical signs | alien hand or limb, orobuccal apraxia, apraxia, ideomotor apraxia, cannot pantomime, limb apraxia, unable to stick out tongue, unable to lick lips, unable to whistle, simultanagnosia, cortical sensory deficit |
| UMN signs | upper motor neuron (UMN) signs, pseudobulbar palsy, inappropriate/pathological laughter, inappropriate/pathological crying, hyperreflexia, positive Hoffmann sign/reflex, positive/extensor/upgoing (not absent/flexor/downgoing) Babinski sign/reflex, positive/extensor/upgoing (not absent/flexor/downgoing) plantar reflex, spasticity |
| LMN signs | lower motor neuron (LMN) signs, fasciculations, tongue atrophy |
| Motor neuron signs | upper motor neuron (UMN) signs, pseudobulbar palsy, inappropriate/pathological laughter, inappropriate/pathological crying, hyperreflexia, positive Hoffmann sign/reflex, positive/extensor/upgoing (not absent/flexor/downgoing) Babinski sign/reflex, positive/extensor/upgoing (not absent/flexor/downgoing) plantar reflex, spasticity, lower motor neuron (LMN) signs, fasciculations, tongue atrophy |
| Dysphagia | swallowing difficulty, aspiration, PEG/feeding tube insertion |
| Any ataxia | ataxia, scanned speech, hypermetric saccades, overshoot, positive or abnormal Romberg sign, dysmetria, hypermetria, abnormal finger to nose, abnormal heel to shin |
| Any amnesia | memory loss//deficit, memory loss/deficit benefited by cues, amnesia |
| Any prosopagnosia | prosopagnosia, unable to answer famous faces |
| Limb rigidity/myoclonus | appendicular (limb) rigidity, limb myoclonus |
| Any hyposmia | hyposmia, loss of smell |
| Any dystonia | dystonia, antecollis, antecollis, retrocollis, clenched fist, hand dystonia, foot dystonia |
| Tremor | shaking, tremor, tremor at rest, postural tremor, kinetic tremor, essential tremor (ET), jaw/chin tremor |
| Any urinary dysfunction | urinary dysfunction, urinary incontinence, urinary catheter |
|  | At least 2 of the following items |
| Autonomic dysfunction | orthostatic hypotension, Any urinary dysfunction, erectile dysfunction (ED), abnormal sweat, cold discolored hands/feet |
|  | At least 3 of the following items |
| Frontal presentation(≥3) | apathy, social withdrawal, abulia, bradyphrenia, change in personality, disinhibition, impulsivity, obsession, compulsivity, hyperorality, abnormally increased appetite, hyperphagia, perseveration, echolalia, palilalia, groping, speaking out of turn, stereotyped behavior, joking/jovial behavior, kleptomania/stealing, cussing behavior, hypersexuality, caught in a scam/fraud, gambling, addiction/abuse, Baker acted, suicidal ideation, criminal behavior, hospitalized in psychiatric ward/asylum, positive grasping reflex, positive snout reflex, positive sucking reflex, positive palmomental reflex, frontal release sign, gegenhalten, paratonia, abnormal Luria test, abnormal fist-edge-palm test, applause sign, abnormal go-no-go test |
| Ataxia | ataxia, scanned speech, hypermetric saccades, overshoot, positive or abnormal Romberg sign, dysmetria, hypermetria, abnormal finger to nose, abnormal heel to shin |
|  | Combination of syndromes[11, 13] |
| Parkinsonism | Bradykinesia & (Rigidity or Rest tremor) |
| CBS (C3, MDS 2017) | Cortical signs & Limb rigidity/myoclonus |
| prob. PSP-RS (MDS 2017) | O1-2 & P1-2 |
| prob. PSP-PGF (MDS 2017) | Akinesia & P1-2 |
| prob. PSP-P (MDS 2017) | O1-2 & Parkinsonism |
| poss. PSP-SL (MDS 2017) | O1-2 & Speech/language disorder |
| poss. PSP-CBS (MDS 2017) | O1-2 & CBS |
| prob. PSP-F (MDS 2017) | O1-2 & Frontal presentation |
| PSP-C | O1-2 & Ataxia |

CBS, Corticobasal syndrome; PSP, progressive supranuclear palsy; PSP-RS, PSP-Richardson’s syndrome; PSP-PGF, PSP-progressive gait freezing; PSP-P, PSP-parkinsonism; PSP-SL, PSP-speech/language disorders; PSP-CBS, PSP-corticobasal syndrome; PSP-F, PSP- frontal presentation; PSP-C, PSP-cerebellar variant; O1-2, level 1 or level 2 of ocular motor dysfunction; P1-2, level 1 or level 2 of postural instability

**Supplementary Table 4 Multivariable Cox proportional-hazards model with subtypes identified by clustering analysis**

| **Covariate** | **Hazard Ratio** | ***P*-value** |
| --- | --- | --- |
| S2 | 0.55 | **<0.01** |
| S3 | 0.56 | **<0.01** |
| S4 | 0.43 | **<0.01** |
| S5 | 0.52 | **<0.01** |
| S6 | 0.37 | **<0.01** |
| Age (years) | 0.99 | **0.03** |
| Sex (Male) | 0.97 | 0.76 |

S1 is used as a reference

**Supplementary Table 5 Multivariable Cox proportional-hazards model with subtypes with early PSP subtypes**

| **Covariate** | **Hazard Ratio** | ***P*-value** |
| --- | --- | --- |
| PSP-RS | 0.63 | **<0.01** |
| PSP-PI | 0.50 | **<0.01** |
| PSP-P | 0.35 | **<0.01** |
| PSP-SL | 0.47 | **<0.01** |
| PSP-F | 0.37 | **<0.01** |
| PSP-OM | 0.49 | **0.01** |
| Misc | 0.42 | **<0.01** |
| Age (years) | 0.99 | **0.04** |
| Sex (Male) | 0.95 | 0.57 |

PSP-PF is used as a reference. F, frontal presentation; Misc, miscellaneous presentations; OM, ocular motor dysfunction; P, parkinsonism; PF, postural and frontal dysfunction; PI, postural instability; PSP, progressive supranuclear palsy; RS, Richardson’s syndrome; SL, speech/language disorder

**Supplementary Table 6 Early clinical features between early PSP subtypes**

|  | **Overall** |  |  |  | **Subtypes** |  |  |  |  | ***P* value** |
| --- | --- | --- | --- | --- | --- | --- | --- | --- | --- | --- |
|  |  | **PSP-PF** | **PSP-RS** | **PSP-PI** | **PSP-P** | **PSP-SL** | **PSP-F** | **PSP-OM** | **Misc** |  |
| ***n*** | 588 | 188 | 68 | 204 | 36 | 29 | 31 | 13 | 19 |  |
| *12 Key features* |  |  |  |  |  |  |  |  |  |  |
| Ocular motor dysfunction | 211/286 (73.8%) | 122/147 (83.0%)^P,PI,RS^ | 68/68 (100.0%)^F,Mi,P,PI,PF,SL^ | 0/36 (0.0%)^F,OM,PF,RS,SL^ | 0/6 (0.0%)^OM,PF,RS^ | 5/9 (55.6%)^PI,RS^ | 3/6 (50.0%)^PI,RS^ | 13/13 (100.0%)^Mi,P,PI^ | 0/1 (0.0%)^OM,RS^ | **<0.01** |
| Postural instability | 460/471 (97.7%) | 188/188 (100.0%)^F,OM,P,SL^ | 68/68 (100.0%)^F,OM,P,SL^ | 204/204 (100.0%)^F,OM,P,SL^ | 0/2 (0.0%)^PI,PF,RS^ | 0/4 (0.0%)^PI,PF,RS^ | 0/4 (0.0%)^PI,PF,RS^ | 0/1 (0.0%)^PI,PF,RS^ | NA | **<0.01** |
| Bradykinesia | 227/237 (95.8%) | 133/135 (98.5%)^SL^ | 37/40 (92.5%)^SL^ | 30/32 (93.8%)^SL^ | 12/12 (100.0%)^SL^ | 2/5 (40.0%)^P,PI,PF,RS^ | 8/8 (100.0%) | 5/5 (100.0%) | NA | **<0.01** |
| Rigidity | 197/252 (78.2%) | 112/135 (83.0%) | 32/42 (76.2%) | 26/38 (68.4%) | 13/15 (86.7%) | 4/7 (57.1%) | 5/7 (71.4%) | 5/7 (71.4%) | 0/1 (0.0%) | 0.17 |
| Rest tremor | 65/140 (46.4%) | 35/78 (44.9%) | 13/23 (56.5%) | 7/20 (35.0%) | 8/10 (80.0%) | 0/1 (0.0%) | 1/4 (25.0%) | 1/3 (33.3%) | 0/1 (0.0%) | 0.23 |
| Akinesia | 66/95 (69.5%) | 38/57 (66.7%) | 9/13 (69.2%) | 8/12 (66.7%) | 4/5 (80.0%) | 2/2 (100.0%) | 2/3 (66.7%) | 2/2 (100.0%) | 1/1 (100.0%) | 0.91 |
| Frontal presentation | 231/285 (81.1%) | 188/188 (100.0%)^OM,Mi,P,PI,RS,SL^ | 0/20 (0.0%)^F,PF,SL^ | 0/20 (0.0%)^F,PF,SL^ | 0/5 (0.0%)^F,PF,SL^ | 12/17 (70.6%)^F,Mi,P,PI,PF,RS^ | 31/31 (100.0%)^OM,Mi,P,PI,RS,SL^ | 0/1 (0.0%)^F,PF^ | 0/3 (0.0%)^F,PF,SL^ | **<0.01** |
| Speech/language disorder | 131/199 (65.8%) | 78/102 (76.5%)^F,P,PI,RS,SL^ | 8/26 (30.8%)^PF,SL^ | 16/33 (48.5%)PF^,SL^ | 0/3 (0.0%)PF^,SL^ | 29/29 (100.0%)^F,OM,Mi,P,PI,PF,RS^ | 0/3 (0.0%)PF^,SL^ | 0/2 (0.0%)^SL^ | 0/1 (0.0%)^SL^ | **<0.01** |
| Cortical signs | 79/120 (65.8%) | 50/71 (70.4%) | 9/17 (52.9%) | 7/16 (43.8%) | 2/3 (66.7%) | 6/6 (100.0%) | 1/3 (33.3%) | 3/3 (100.0%) | 1/1 (100.0%) | 0.1 |
| Limb rigidity/myoclonus | 24/35 (68.6%) | 15/23 (65.2%) | 2/4 (50.0%) | 5/5 (100.0%) | 0/1 (0.0%) | NA | NA | 2/2 (100.0%) | NA | 0.19 |
| Motor neuron signs | 135/270 (50.0%) | 81/135 (60.0%) | 20/45 (44.4%) | 24/58 (41.4%) | 1/6 (16.7%) | 1/10 (10.0%) | 4/6 (66.7%) | 2/4 (50.0%) | 2/6 (33.3%) | **0.01** |
| Ataxia | 16/119 (13.4%) | 12/71 (16.9%) | 3/21 (14.3%) | 1/20 (5.0%) | 0/1 (0.0%) | 0/2 (0.0%) | 0/2 (0.0%) | 0/1 (0.0%) | 0/1 (0.0%) | 0.88 |
| *Syndromes based on previous literature* |  |  |  |  |  |  |  |  |  |  |
| prob. PSP-RS (MDS 2017) | 187/257 (72.8%) | 121/145 (83.4%)^F,OM,P,PI,RS,SL^ | 66/67 (98.5%)^F,OM,P,PI,PF,SL^ | 0/36 (0.0%)^PF,RS^ | 0/3 (0.0%)^PF,RS^ | 0/3 (0.0%)^PF,RS^ | 0/2 (0.0%)^PF,RS^ | 0/1 (0.0%)^PF,RS^ | NA | **<0.01** |
| prob. PSP-PGF (MDS 2017) | 40/77 (51.9%) | 29/52 (55.8%) | 8/13 (61.5%) | 0/3 (0.0%) | 0/3 (0.0%) | 1/2 (50.0%) | 0/2 (0.0%) | 2/2 (100.0%) | NA | 0.08 |
| prob. PSP-P (MDS 2017) | 122/171 (71.3%) | 87/115 (75.7%)^P,PI^ | 29/35 (82.9%)P,PI | 0/9 (0.0%)^OM,PF,RS,SL^ | 0/4 (0.0%)^PF,RS^ | 2/2 (100.0%)^PI^ | 0/2 (0.0%) | 4/4 (100.0%)^PI^ | NA | **<0.01** |
| Parkinsonism | 168/197 (85.3%) | 104/121 (86.0%) | 30/35 (85.7%) | 18/22 (81.8%) | 6/6 (100.0%) | 2/3 (66.7%) | 4/6 (66.7%) | 4/4 (100.0%) | NA | 0.61 |
| poss. PSP-SL (MDS 2017) | 75/152 (49.3%) | 62/92 (67.4%)^PI,RS^ | 8/26 (30.8%)^PF^ | 0/19 (0.0%)PF^,SL^ | 0/2 (0.0%) | 5/9 (55.6%)^PI^ | 0/2 (0.0%) | 0/2 (0.0%) | NA | **<0.01** |
| poss. PSP-CBS (MDS 2017) | 2/15 (13.3%) | 2/13 (15.4%) | 0/1 (0.0%) | NA | 0/1 (0.0%) | NA | NA | NA | NA | 0.84 |
| CBS (C3, MDS 2017) | 3/16 (18.8%) | 2/13 (15.4%) | 0/1 (0.0%) | NA | 0/1 (0.0%) | NA | NA | 1/1 (100.0%) | NA | 0.18 |
| prob. PSP-F (MDS 2017) | 43/92 (46.7%) | 42/82 (51.2%) | 0/6 (0.0%) | 0/1 (0.0%) | NA | 1/2 (50.0%) | 0/1 (0.0%) | NA | NA | 0.1 |
| PSP-C | 13/103 (12.6%) | 10/65 (15.4%) | 3/21 (14.3%) | 0/13 (0.0%) | 0/1 (0.0%) | 0/1 (0.0%) | 0/1 (0.0%) | 0/1 (0.0%) | NA | 0.81 |
| *Grouped features* |  |  |  |  |  |  |  |  |  |  |
| O1-2 (MDS 2017) | 207/282 (73.4%) | 121/145 (83.4%)^P,PI,RS^ | 66/67 (98.5%)^F,Mi,P,PI,PF,SL^ | 0/36 (0.0%)^F,OM,PF,RS,SL^ | 0/6 (0.0%)^OM,PF,RS^ | 5/9 (55.6%)^PI,RS^ | 3/6 (50.0%)^PI,RS^ | 12/12 (100.0%)^Mi,P,PI^ | 0/1 (0.0%)^OM,RS^ | **<0.01** |
| P1-2 (MDS 2017) | 461/473 (97.5%) | 188/188 (100.0%)^F,OM,P,SL^ | 68/68 (100.0%)^F,OM,P,SL^ | 204/204 (100.0%)^F,OM,P,SL^ | 1/3 (33.3%)^PI,PF,RS^ | 0/4 (0.0%)^PI,PF,RS^ | 0/5 (0.0%)^PI,PF,RS^ | 0/1 (0.0%)^PI,PF,RS^ | NA | **<0.01** |
| Any parkinsonian gait | 88/103 (85.4%) | 49/56 (87.5%) | 21/23 (91.3%) | 12/14 (85.7%) | 3/4 (75.0%) | 0/1 (0.0%) | 1/2 (50.0%) | 2/3 (66.7%) | NA | 0.13 |
| Any abnormal posture | 81/89 (91.0%) | 47/51 (92.2%) | 16/18 (88.9%) | 11/11 (100.0%) | 3/3 (100.0%) | 1/2 (50.0%) | 2/3 (66.7%) | 1/1 (100.0%) | NA | 0.24 |
| UMN signs | 128/250 (51.2%) | 77/125 (61.6%) | 20/44 (45.5%) | 21/54 (38.9%) | 1/5 (20.0%) | 1/8 (12.5%) | 4/5 (80.0%) | 2/4 (50.0%) | 2/5 (40.0%) | **0.01** |
| LMN signs | 16/125 (12.8%) | 10/71 (14.1%) | 1/15 (6.7%) | 3/23 (13.0%) | 0/2 (0.0%) | 0/8 (0.0%) | 1/2 (50.0%) | 1/2 (50.0%) | 0/2 (0.0%) | 0.4 |
| Dysphagia | 145/215 (67.4%) | 89/122 (73.0%) | 17/30 (56.7%) | 27/41 (65.9%) | 4/6 (66.7%) | 2/5 (40.0%) | 5/7 (71.4%) | 1/3 (33.3%) | 0/1 (0.0%) | 0.27 |
| Frontal presentation(≥3) | 58/109 (53.2%) | 53/92 (57.6%)^RS^ | 0/6 (0.0%)^F,PF^ | 0/2 (0.0%) | NA | 3/7 (42.9%) | 2/2 (100.0%)^RS^ | NA | NA | **0.02** |
| Any ataxia | 138/267 (51.7%) | 78/136 (57.4%) | 21/45 (46.7%) | 26/56 (46.4%) | 3/7 (42.9%) | 1/9 (11.1%) | 2/4 (50.0%) | 3/3 (100.0%) | 4/7 (57.1%) | 0.11 |
| Any amnesia | 270/313 (86.3%) | 138/158 (87.3%) | 39/45 (86.7%) | 56/66 (84.8%) | 4/7 (57.1%) | 16/18 (88.9%) | 9/11 (81.8%) | 4/4 (100.0%) | 4/4 (100.0%) | 0.45 |
| Any prosopagnosia | 0/1 (0.0%) | 0/1 (0.0%) | NA | NA | NA | NA | NA | NA | NA | 1 |
| Any hyposmia | 25/56 (44.6%) | 17/39 (43.6%) | 2/6 (33.3%) | 3/5 (60.0%) | 0/1 (0.0%) | 1/3 (33.3%) | 2/2 (100.0%) | NA | NA | 0.51 |
| Any dystonia | 62/77 (80.5%) | 33/44 (75.0%) | 12/13 (92.3%) | 9/10 (90.0%) | 0/1 (0.0%) | 1/2 (50.0%) | 3/3 (100.0%) | 4/4 (100.0%) | NA | 0.14 |
| Autonomic dysfunction | 38/85 (44.7%) | 29/63 (46.0%) | 5/9 (55.6%) | 3/8 (37.5%) | 1/3 (33.3%) | 0/1 (0.0%) | 0/1 (0.0%) | NA | NA | 0.79 |
| Any urinary dysfunction | 148/215 (68.8%) | 79/116 (68.1%) | 22/33 (66.7%) | 26/38 (68.4%) | 6/7 (85.7%) | 7/10 (70.0%) | 5/7 (71.4%) | 2/2 (100.0%) | 1/2 (50.0%) | 0.94 |
| *Individual features* |  |  |  |  |  |  |  |  |  |  |
| hypomimia | 44/48 (91.7%) | 29/30 (96.7%)F | 8/10 (80.0%) | 5/5 (100.0%) | 2/2 (100.0%) | NA | 0/1 (0.0%)^PF^ | NA | NA | **<0.01** |
| masked face | 24/25 (96.0%) | 14/14 (100.0%)^PI^ | 5/5 (100.0%) | 1/2 (50.0%)^PF^ | 1/1 (100.0%) | NA | 2/2 (100.0%) | 1/1 (100.0%) | NA | **0.04** |
| bradykinesia | 154/166 (92.8%) | 85/91 (93.4%) | 29/31 (93.5%) | 20/23 (87.0%) | 9/9 (100.0%) | 1/2 (50.0%) | 6/6 (100.0%) | 4/4 (100.0%) | NA | 0.23 |
| slow movement | 168/171 (98.2%) | 112/112 (100.0%)^SL^ | 25/25 (100.0%)^SL^ | 16/17 (94.1%) | 5/5 (100.0%) | 2/4 (50.0%)^PF,RS^ | 4/4 (100.0%) | 4/4 (100.0%) | NA | **<0.01** |
| decreased arm swing | 115/145 (79.3%) | 68/86 (79.1%) | 23/25 (92.0%) | 14/19 (73.7%) | 3/6 (50.0%) | 2/4 (50.0%) | 3/3 (100.0%) | 2/2 (100.0%) | NA | 0.15 |
| micrographia | 45/55 (81.8%) | 26/33 (78.8%) | 6/7 (85.7%) | 10/11 (90.9%) | 2/2 (100.0%) | NA | NA | 1/1 (100.0%) | 0/1 (0.0%) | 0.3 |
| small handwriting | 63/75 (84.0%) | 33/42 (78.6%) | 7/9 (77.8%) | 14/15 (93.3%) | 4/4 (100.0%) | NA | 4/4 (100.0%) | 1/1 (100.0%) | NA | 0.57 |
| akinesia | 23/25 (92.0%) | 12/13 (92.3%) | 2/2 (100.0%) | 3/3 (100.0%) | 3/4 (75.0%) | NA | 1/1 (100.0%) | 2/2 (100.0%) | NA | 0.81 |
| no spontaneous movements | 20/31 (64.5%) | 12/19 (63.2%) | 2/3 (66.7%) | 1/4 (25.0%) | 2/2 (100.0%) | 2/2 (100.0%) | 1/1 (100.0%) | NA | NA | 0.36 |
| gait freezing | 27/45 (60.0%) | 16/28 (57.1%) | 5/8 (62.5%) | 5/6 (83.3%) | 0/1 (0.0%) | NA | 0/1 (0.0%) | NA | 1/1 (100.0%) | 0.4 |
| magnetic gait | 2/3 (66.7%) | 2/3 (66.7%) | NA | NA | NA | NA | NA | NA | NA | 1 |
| rigidity | 170/197 (86.3%) | 100/113 (88.5%)SL | 27/31 (87.1%) | 24/27 (88.9%) | 10/11 (90.9%) | 1/4 (25.0%)RS | 4/5 (80.0%) | 4/6 (66.7%) | NA | 0.02 |
| axial rigidity | 42/51 (82.4%) | 34/41 (82.9%) | 5/6 (83.3%) | 3/3 (100.0%) | 0/1 (0.0%) | NA | NA | NA | NA | 0.15 |
| nuchal (neck) rigidity | 54/63 (85.7%) | 40/48 (83.3%) | 10/11 (90.9%) | 3/3 (100.0%) | 1/1 (100.0%) | NA | NA | NA | NA | 0.77 |
| appendicular (limb) rigidity | 23/34 (67.6%) | 15/23 (65.2%) | 2/4 (50.0%) | 5/5 (100.0%) | 0/1 (0.0%) | NA | NA | 1/1 (100.0%) | NA | 0.23 |
| cogwheel rigidity | 75/113 (66.4%) | 38/62 (61.3%) | 14/18 (77.8%) | 8/13 (61.5%) | 7/8 (87.5%) | 3/5 (60.0%) | 3/4 (75.0%) | 2/3 (66.7%) | NA | 0.71 |
| stiffness | 89/109 (81.7%) | 47/61 (77.0%) | 11/15 (73.3%) | 19/20 (95.0%) | 6/6 (100.0%) | 1/1 (100.0%) | 3/4 (75.0%) | 2/2 (100.0%) | NA | 0.41 |
| increased muscle tone | 38/86 (44.2%) | 28/48 (58.3%) | 4/16 (25.0%) | 2/12 (16.7%) | 2/3 (66.7%) | 1/4 (25.0%) | 0/1 (0.0%) | 1/1 (100.0%) | 0/1 (0.0%) | 0.05 |
| the fact patient fell, not risk of fall | 191/191 (100.0%) | 89/89 (100.0%) | 23/23 (100.0%) | 59/59 (100.0%) | 3/3 (100.0%) | 5/5 (100.0%) | 4/4 (100.0%) | 3/3 (100.0%) | 5/5 (100.0%) | 1 |
| multiple/frequent falls | 458/469 (97.7%) | 188/188 (100.0%)^F,OM,P,SL^ | 67/67 (100.0%)^F,OM,P,SL^ | 203/203 (100.0%)^F,OM,P,SL^ | 0/2 (0.0%)^PI,PF,RS^ | 0/4 (0.0%)^PI,PF,RS^ | 0/4 (0.0%)^PI,PF,RS^ | 0/1 (0.0%)^PI,PF,RS^ | NA | **<0.01** |
| falling forwards | 68/69 (98.6%) | 34/34 (100.0%)^P^ | 12/12 (100.0%)^P^ | 22/22 (100.0%)^P^ | 0/1 (0.0%)^PI,PF,RS^ | NA | NA | NA | NA | **<0.01** |
| positive pull test | 44/66 (66.7%) | 29/46 (63.0%) | 10/12 (83.3%) | 4/5 (80.0%) | 1/1 (100.0%) | 0/1 (0.0%) | 0/1 (0.0%) | NA | NA | 0.25 |
| falling backwards | 130/131 (99.2%) | 65/65 (100.0%)^P^ | 23/23 (100.0%)^P^ | 42/42 (100.0%)P | 0/1 (0.0%)^PI,PF,RS^ | NA | NA | NA | NA | **<0.01** |
| imbalance | 141/144 (97.9%) | 74/74 (100.0%)^F,SL^ | 24/24 (100.0%)^F,SL^ | 38/39 (97.4%)SL | 1/1 (100.0%) | 1/2 (50.0%)^PI,PF,RS^ | 2/3 (66.7%)^PF,RS^ | 1/1 (100.0%) | NA | **<0.01** |
| shuffling gait | 68/80 (85.0%) | 38/43 (88.4%) | 14/15 (93.3%) | 10/12 (83.3%) | 3/4 (75.0%) | 0/1 (0.0%) | 1/2 (50.0%) | 2/3 (66.7%) | NA | 0.13 |
| festinating gait | 13/22 (59.1%) | 9/14 (64.3%) | 2/4 (50.0%) | 1/3 (33.3%) | NA | NA | NA | 1/1 (100.0%) | NA | 0.61 |
| turning en bloc | 24/27 (88.9%) | 16/17 (94.1%) | 7/8 (87.5%) | 1/2 (50.0%) | NA | NA | NA | NA | NA | 0.17 |
| stooped posture | 74/79 (93.7%) | 45/47 (95.7%) | 15/16 (93.8%) | 8/8 (100.0%) | 3/3 (100.0%) | 1/2 (50.0%) | 1/2 (50.0%) | 1/1 (100.0%) | NA | **0.03** |
| pisa sign | 1/1 (100.0%) | 1/1 (100.0%) | NA | NA | NA | NA | NA | NA | NA | 1 |
| camptocormia | 1/1 (100.0%) | 1/1 (100.0%) | NA | NA | NA | NA | NA | NA | NA | 1 |
| scoliosis | 15/20 (75.0%) | 7/11 (63.6%) | 2/3 (66.7%) | 5/5 (100.0%) | NA | NA | 1/1 (100.0%) | NA | NA | 0.41 |
| memory loss//deficit | 265/309 (85.8%) | 138/158 (87.3%) | 38/44 (86.4%) | 53/64 (82.8%) | 4/7 (57.1%) | 16/18 (88.9%) | 9/11 (81.8%) | 4/4 (100.0%) | 3/3 (100.0%) | 0.44 |
| memory loss/deficit benefited by cues | 27/28 (96.4%) | 17/18 (94.4%) | 2/2 (100.0%) | 4/4 (100.0%) | NA | 4/4 (100.0%) | NA | NA | NA | 0.9 |
| amnesia | 15/20 (75.0%) | 7/12 (58.3%) | 1/1 (100.0%) | 5/5 (100.0%) | NA | 1/1 (100.0%) | NA | NA | 1/1 (100.0%) | 0.35 |
| word finding difficulty | 92/99 (92.9%) | 53/57 (93.0%) | 9/11 (81.8%) | 13/13 (100.0%) | NA | 15/15 (100.0%) | 2/3 (66.7%) | NA | NA | 0.12 |
| disorientation | 64/250 (25.6%) | 40/131 (30.5%) | 8/36 (22.2%) | 8/53 (15.1%) | 1/5 (20.0%) | 5/13 (38.5%) | 0/5 (0.0%) | 1/3 (33.3%) | 1/4 (25.0%) | 0.33 |
| lost name | 3/5 (60.0%) | 1/3 (33.3%) | NA | 1/1 (100.0%) | NA | NA | 1/1 (100.0%) | NA | NA | 0.33 |
| lost place | 2/4 (50.0%) | 1/2 (50.0%) | NA | 0/1 (0.0%) | NA | 1/1 (100.0%) | NA | NA | NA | 0.37 |
| impaired serial sevens | 27/33 (81.8%) | 16/20 (80.0%) | 3/3 (100.0%) | 4/4 (100.0%) | 1/2 (50.0%) | 2/3 (66.7%) | 1/1 (100.0%) | NA | NA | 0.6 |
| abnormal clock test | 11/14 (78.6%) | 10/11 (90.9%) | NA | 1/2 (50.0%) | NA | 0/1 (0.0%) | NA | NA | NA | 0.06 |
| impaired spelling WORLD backwards | 15/25 (60.0%) | 8/14 (57.1%) | 3/5 (60.0%) | NA | 0/1 (0.0%) | 2/3 (66.7%) | 2/2 (100.0%) | NA | NA | 0.57 |
| wander | 8/8 (100.0%) | 4/4 (100.0%) | 1/1 (100.0%) | NA | NA | NA | 2/2 (100.0%) | 1/1 (100.0%) | NA | 1 |
| gets lost | 16/34 (47.1%) | 9/21 (42.9%) | 1/2 (50.0%) | 3/6 (50.0%) | 1/1 (100.0%) | 1/3 (33.3%) | 1/1 (100.0%) | NA | NA | 0.75 |
| prosopagnosia | 0/1 (0.0%) | 0/1 (0.0%) | NA | NA | NA | NA | NA | NA | NA | 1 |
| fluctuation of neuropsychiatric symptoms | 22/32 (68.8%) | 19/27 (70.4%) | 1/1 (100.0%) | 0/1 (0.0%) | NA | 1/1 (100.0%) | NA | 1/1 (100.0%) | 0/1 (0.0%) | 0.33 |
| sundowning | 1/1 (100.0%) | 1/1 (100.0%) | NA | NA | NA | NA | NA | NA | NA | 1 |
| hallucinations | 29/133 (21.8%) | 19/89 (21.3%) | 5/17 (29.4%) | 3/11 (27.3%) | NA | 0/9 (0.0%) | 0/3 (0.0%) | 1/2 (50.0%) | 1/2 (50.0%) | 0.42 |
| visual hallucinations | 25/131 (19.1%) | 16/89 (18.0%) | 5/17 (29.4%) | 2/9 (22.2%) | NA | 0/9 (0.0%) | 0/3 (0.0%) | 1/2 (50.0%) | 1/2 (50.0%) | 0.36 |
| paranoid delusions | 12/20 (60.0%) | 7/15 (46.7%) | NA | 1/1 (100.0%) | NA | NA | 3/3 (100.0%) | NA | 1/1 (100.0%) | 0.22 |
| apathy | 33/42 (78.6%) | 32/39 (82.1%)^RS^ | 0/2 (0.0%)^PF^ | NA | NA | NA | 1/1 (100.0%) | NA | NA | **0.02** |
| social withdrawal | 45/45 (100.0%) | 37/37 (100.0%) | NA | NA | NA | 4/4 (100.0%) | 4/4 (100.0%) | NA | NA | 1 |
| bradyphrenia | 41/44 (93.2%) | 36/38 (94.7%)^RS^ | 0/1 (0.0%)^PF^ | NA | NA | 2/2 (100.0%) | 3/3 (100.0%) | NA | NA | **<0.01** |
| change in personality | 80/97 (82.5%) | 60/69 (87.0%)^PI,SL^ | 0/1 (0.0%)^F^ | 0/3 (0.0%)^F,PF^ | 0/1 (0.0%)^F^ | 2/5 (40.0%)^F,PF^ | 18/18 (100.0%)^P,PI,RS,SL^ | NA | NA | **<0.01** |
| disinhibition | 16/21 (76.2%) | 16/21 (76.2%) | NA | NA | NA | NA | NA | NA | NA | 1 |
| impulsivity | 35/43 (81.4%) | 32/39 (82.1%) | NA | NA | NA | 1/2 (50.0%) | 2/2 (100.0%) | NA | NA | 0.41 |
| obsession | 6/12 (50.0%) | 5/10 (50.0%) | NA | NA | NA | 1/2 (50.0%) | NA | NA | NA | 1 |
| compulsivity | 7/10 (70.0%) | 5/8 (62.5%) | NA | NA | NA | 1/1 (100.0%) | 1/1 (100.0%) | NA | NA | 0.59 |
| abnormally increased appetite | 6/99 (6.1%) | 5/62 (8.1%) | 0/11 (0.0%) | 0/11 (0.0%) | 0/4 (0.0%) | 0/6 (0.0%) | 1/4 (25.0%) | 0/1 (0.0%) | NA | 0.53 |
| perseveration | 24/30 (80.0%) | 22/26 (84.6%) | 0/1 (0.0%) | NA | NA | 2/3 (66.7%) | NA | NA | NA | 0.1 |
| echolalia | 6/6 (100.0%) | 5/5 (100.0%) | NA | NA | NA | 1/1 (100.0%) | NA | NA | NA | 1 |
| palilalia | 5/6 (83.3%) | 3/3 (100.0%) | 0/1 (0.0%) | NA | NA | 1/1 (100.0%) | 1/1 (100.0%) | NA | NA | 0.11 |
| groping | 2/2 (100.0%) | 1/1 (100.0%) | NA | NA | NA | 1/1 (100.0%) | NA | NA | NA | 1 |
| speaking out of turn | 5/5 (100.0%) | 5/5 (100.0%) | NA | NA | NA | NA | NA | NA | NA | 1 |
| stereotyped behavior | 0/4 (0.0%) | 0/4 (0.0%) | NA | NA | NA | NA | NA | NA | NA | 1 |
| joking/jovial behavior | 6/7 (85.7%) | 5/6 (83.3%) | NA | NA | NA | 1/1 (100.0%) | NA | NA | NA | 0.66 |
| cussing behavior | 1/1 (100.0%) | 1/1 (100.0%) | NA | NA | NA | NA | NA | NA | NA | 1 |
| hypersexuality | 5/13 (38.5%) | 4/9 (44.4%) | NA | 0/2 (0.0%) | NA | 0/1 (0.0%) | 1/1 (100.0%) | NA | NA | 0.31 |
| gambling | 2/3 (66.7%) | 2/3 (66.7%) | NA | NA | NA | NA | NA | NA | NA | 1 |
| addiction/abuse | 28/58 (48.3%) | 23/44 (52.3%) | 0/2 (0.0%) | 0/2 (0.0%) | NA | 3/6 (50.0%) | 2/2 (100.0%) | NA | 0/2 (0.0%) | 0.15 |
| suicidal ideation | 17/59 (28.8%) | 14/41 (34.1%) | 0/4 (0.0%) | 0/5 (0.0%) | 0/1 (0.0%) | 1/6 (16.7%) | 2/2 (100.0%) | NA | NA | 0.08 |
| criminal behavior | 1/3 (33.3%) | 1/3 (33.3%) | NA | NA | NA | NA | NA | NA | NA | 1 |
| hospitalized in psychiatric ward/asylum | 10/15 (66.7%) | 9/14 (64.3%) | NA | NA | NA | NA | 1/1 (100.0%) | NA | NA | 0.46 |
| positive grasping reflex | 12/27 (44.4%) | 11/21 (52.4%) | 0/3 (0.0%) | 0/1 (0.0%) | NA | 0/1 (0.0%) | 1/1 (100.0%) | NA | NA | 0.22 |
| positive snout reflex | 17/25 (68.0%) | 15/20 (75.0%) | 0/2 (0.0%) | 0/1 (0.0%) | NA | NA | 2/2 (100.0%) | NA | NA | 0.05 |
| positive sucking reflex | 1/1 (100.0%) | NA | NA | NA | NA | NA | 1/1 (100.0%) | NA | NA | 1 |
| positive palmomental reflex | 18/33 (54.5%) | 16/26 (61.5%) | 0/3 (0.0%) | 0/1 (0.0%) | NA | 1/1 (100.0%) | 1/1 (100.0%) | NA | 0/1 (0.0%) | 0.15 |
| frontal release sign | 13/24 (54.2%) | 12/17 (70.6%) | 0/3 (0.0%) | 0/2 (0.0%) | NA | 0/1 (0.0%) | 1/1 (100.0%) | NA | NA | **0.04** |
| gegenhalten | 5/6 (83.3%) | 5/5 (100.0%)^SL^ | NA | NA | NA | 0/1 (0.0%)^PF^ | NA | NA | NA | **0.01** |
| paratonia | 12/12 (100.0%) | 12/12 (100.0%) | NA | NA | NA | NA | NA | NA | NA | 1 |
| abnormal Luria test | 7/8 (87.5%) | 7/8 (87.5%) | NA | NA | NA | NA | NA | NA | NA | 1 |
| applause sign | 14/21 (66.7%) | 14/18 (77.8%)^RS^ | 0/3 (0.0%)^PF^ | NA | NA | NA | NA | NA | NA | **<0.01** |
| abnormal go-no-go test | 3/3 (100.0%) | 3/3 (100.0%) | NA | NA | NA | NA | NA | NA | NA | 1 |
| stopped driving | 31/35 (88.6%) | 24/27 (88.9%) | NA | 5/6 (83.3%) | NA | 1/1 (100.0%) | 1/1 (100.0%) | NA | NA | 0.94 |
| car accident | 28/30 (93.3%) | 12/14 (85.7%) | 4/4 (100.0%) | 7/7 (100.0%) | NA | 3/3 (100.0%) | NA | 2/2 (100.0%) | NA | 0.65 |
| loss of appetite | 30/100 (30.0%) | 20/63 (31.7%) | 3/11 (27.3%) | 4/11 (36.4%) | 0/4 (0.0%) | 1/6 (16.7%) | 2/4 (50.0%) | 0/1 (0.0%) | NA | 0.71 |
| cries a lot | 40/52 (76.9%) | 29/34 (85.3%) | 3/7 (42.9%) | 4/5 (80.0%) | NA | 1/1 (100.0%) | 2/2 (100.0%) | 1/2 (50.0%) | 0/1 (0.0%) | 0.09 |
| aphasia/primary progressive aphasia (PPA) | 49/90 (54.4%) | 27/49 (55.1%) | 2/7 (28.6%) | 5/13 (38.5%) | 0/1 (0.0%) | 15/17 (88.2%) | 0/1 (0.0%) | 0/1 (0.0%) | 0/1 (0.0%) | **0.03** |
| agrammatic aphasia | 1/2 (50.0%) | NA | NA | 1/1 (100.0%) | NA | 0/1 (0.0%) | NA | NA | NA | 0.16 |
| reduced fluency | 68/127 (53.5%) | 43/69 (62.3%) | 4/16 (25.0%)^SL^ | 9/23 (39.1%)^SL^ | 0/2 (0.0%)^SL^ | 12/13 (92.3%)^F,P,PI,RS^ | 0/3 (0.0%)^SL^ | 0/1 (0.0%) | NA | **<0.01** |
| apraxia of speech | 13/18 (72.2%) | 4/6 (66.7%) | 0/2 (0.0%)^SL^ | 1/2 (50.0%) | NA | 8/8 (100.0%)^RS^ | NA | NA | NA | **0.03** |
| abnormal articulation/pronunciation | 36/55 (65.5%) | 20/30 (66.7%) | 2/5 (40.0%) | 6/10 (60.0%) | 0/2 (0.0%)SL | 8/8 (100.0%)P | NA | NA | NA | 0.05 |
| spasmodic speech/dysphonia | 1/1 (100.0%) | 1/1 (100.0%) | NA | NA | NA | NA | NA | NA | NA | 1 |
| mutism | 11/15 (73.3%) | 7/10 (70.0%) | 3/4 (75.0%) | NA | 1/1 (100.0%) | NA | NA | NA | NA | 0.81 |
| alien hand or limb | 5/15 (33.3%) | 4/13 (30.8%) | NA | 0/1 (0.0%) | NA | NA | NA | 1/1 (100.0%) | NA | 0.28 |
| apraxia | 71/107 (66.4%) | 46/65 (70.8%) | 7/15 (46.7%) | 6/13 (46.2%) | 2/2 (100.0%) | 6/6 (100.0%) | 1/3 (33.3%) | 2/2 (100.0%) | 1/1 (100.0%) | 0.08 |
| ideomotor apraxia | 6/12 (50.0%) | 6/10 (60.0%) | NA | NA | NA | 0/2 (0.0%) | NA | NA | NA | 0.12 |
| cannot pantomime | 1/1 (100.0%) | NA | 1/1 (100.0%) | NA | NA | NA | NA | NA | NA | 1 |
| limb apraxia | 8/16 (50.0%) | 7/10 (70.0%) | 0/1 (0.0%) | 0/1 (0.0%) | NA | 0/3 (0.0%) | NA | 1/1 (100.0%) | NA | 0.11 |
| unable to lick lips | 0/1 (0.0%) | 0/1 (0.0%) | NA | NA | NA | NA | NA | NA | NA | 1 |
| unable to whistle | 2/3 (66.7%) | NA | 0/1 (0.0%) | 1/1 (100.0%) | NA | 1/1 (100.0%) | NA | NA | NA | 0.22 |
| simultanagnosia | 0/4 (0.0%) | 0/3 (0.0%) | NA | 0/1 (0.0%) | NA | NA | NA | NA | NA | 1 |
| cortical sensory deficit | 4/19 (21.1%) | 3/14 (21.4%) | 1/1 (100.0%) | 0/1 (0.0%) | 0/2 (0.0%) | NA | NA | 0/1 (0.0%) | NA | 0.31 |
| REM sleep behavior disorder (RBD) | 36/76 (47.4%) | 26/53 (49.1%) | 2/9 (22.2%) | 5/6 (83.3%) | NA | 0/4 (0.0%) | 2/3 (66.7%) | 1/1 (100.0%) | NA | 0.06 |
| hyposmia | 3/4 (75.0%) | 2/2 (100.0%) | 0/1 (0.0%) | 1/1 (100.0%) | NA | NA | NA | NA | NA | 0.14 |
| loss of smell | 23/54 (42.6%) | 16/39 (41.0%) | 2/5 (40.0%) | 2/4 (50.0%) | 0/1 (0.0%) | 1/3 (33.3%) | 2/2 (100.0%) | NA | NA | 0.6 |
| restricted eye movement/gaze palsy | 189/265 (71.3%) | 114/137 (83.2%)^P,PI^ | 60/63 (95.2%)^F,OM,Mi,P,PI,SL^ | 0/33 (0.0%)^F,OM,PF,RS,SL^ | 0/5 (0.0%)^PF,RS^ | 4/8 (50.0%)^PI,RS^ | 3/6 (50.0%)^PI,RS^ | 8/12 (66.7%)^PI,RS^ | 0/1 (0.0%)^RS^ | **<0.01** |
| vertical hypometric saccades | 17/19 (89.5%) | 11/13 (84.6%) | 5/5 (100.0%) | NA | NA | NA | NA | 1/1 (100.0%) | NA | 0.6 |
| downgaze palsy | 30/33 (90.9%) | 22/24 (91.7%)^F^ | 7/7 (100.0%)^F^ | NA | NA | NA | 0/1 (0.0%)^PF,RS^ | 1/1 (100.0%) | NA | **0.01** |
| slow saccades | 106/128 (82.8%) | 81/93 (87.1%)^PI^ | 18/22 (81.8%)^PI^ | 0/5 (0.0%)^OM,PF,RS,SL^ | 0/1 (0.0%) | 3/3 (100.0%)^PI^ | NA | 4/4 (100.0%)^PI^ | NA | **<0.01** |
| eyelid opening apraxia | 6/13 (46.2%) | 5/9 (55.6%) | 0/3 (0.0%) | NA | NA | NA | NA | 1/1 (100.0%) | NA | 0.13 |
| blepharospasm | 10/16 (62.5%) | 7/11 (63.6%) | 3/5 (60.0%) | NA | NA | NA | NA | NA | NA | 0.89 |
| photophobia | 9/15 (60.0%) | 6/9 (66.7%) | 3/4 (75.0%) | 0/1 (0.0%) | NA | NA | 0/1 (0.0%) | NA | NA | 0.32 |
| ataxia | 92/137 (67.2%) | 51/73 (69.9%) | 14/23 (60.9%) | 20/27 (74.1%) | 1/2 (50.0%) | 0/4 (0.0%) | 1/2 (50.0%) | 1/1 (100.0%) | 4/5 (80.0%) | 0.15 |
| hypermetric saccades | 0/1 (0.0%) | 0/1 (0.0%) | NA | NA | NA | NA | NA | NA | NA | 1 |
| saccadic hypometria | 16/18 (88.9%) | 11/13 (84.6%) | 5/5 (100.0%) | NA | NA | NA | NA | NA | NA | 0.35 |
| overshoot | 2/6 (33.3%) | 2/4 (50.0%) | 0/2 (0.0%) | NA | NA | NA | NA | NA | NA | 0.22 |
| scanned speech | 1/1 (100.0%) | 1/1 (100.0%) | NA | NA | NA | NA | NA | NA | NA | 1 |
| dysmetria | 22/71 (31.0%) | 13/43 (30.2%) | 5/14 (35.7%) | 3/10 (30.0%) | NA | 0/3 (0.0%) | 1/1 (100.0%) | NA | NA | 0.44 |
| hypermetria | 0/1 (0.0%) | 0/1 (0.0%) | NA | NA | NA | NA | NA | NA | NA | 1 |
| abnormal finger to nose | 36/180 (20.0%) | 27/98 (27.6%) | 3/28 (10.7%)^OM^ | 3/36 (8.3%)^OM^ | 1/6 (16.7%) | 0/6 (0.0%) | 0/2 (0.0%) | 2/2 (100.0%)^PI,RS^ | 0/2 (0.0%) | **<0.01** |
| abnormal heel to shin | 13/111 (11.7%) | 7/60 (11.7%) | 4/19 (21.1%) | 1/24 (4.2%) | NA | 0/3 (0.0%) | 0/1 (0.0%) | 1/2 (50.0%) | 0/2 (0.0%) | 0.36 |
| positive or abnormal Romberg sign | 42/132 (31.8%) | 26/73 (35.6%) | 8/23 (34.8%) | 6/25 (24.0%) | 1/2 (50.0%) | 1/3 (33.3%) | 0/2 (0.0%) | 0/2 (0.0%) | 0/2 (0.0%) | 0.73 |
| square-wave jerks | 44/49 (89.8%) | 35/40 (87.5%) | 7/7 (100.0%) | NA | NA | NA | 1/1 (100.0%) | 1/1 (100.0%) | NA | 0.74 |
| nystagmus | 59/148 (39.9%) | 40/81 (49.4%)^PI^ | 14/25 (56.0%)^PI^ | 3/28 (10.7%)^PF,RS^ | 0/4 (0.0%) | 1/6 (16.7%) | 1/2 (50.0%) | 0/2 (0.0%) | NA | **<0.01** |
| dysdiadochokinesis | 10/23 (43.5%) | 5/13 (38.5%) | 2/6 (33.3%) | 2/3 (66.7%) | NA | NA | NA | 1/1 (100.0%) | NA | 0.5 |
| unable to walk a line | 13/16 (81.2%) | 7/8 (87.5%) | 1/2 (50.0%) | 3/4 (75.0%) | 1/1 (100.0%) | 1/1 (100.0%) | NA | NA | NA | 0.73 |
| unable to perform tandem gait | 71/109 (65.1%) | 40/54 (74.1%) | 13/18 (72.2%) | 14/27 (51.9%) | 1/2 (50.0%) | 2/2 (100.0%) | 1/3 (33.3%) | 0/1 (0.0%) | 0/2 (0.0%) | 0.08 |
| involuntary movement | 55/67 (82.1%) | 31/38 (81.6%) | 7/9 (77.8%) | 9/11 (81.8%) | 3/3 (100.0%) | 2/3 (66.7%) | 2/2 (100.0%) | 1/1 (100.0%) | NA | 0.93 |
| chorea/writhing | 6/13 (46.2%) | 5/11 (45.5%) | 0/1 (0.0%) | 1/1 (100.0%) | NA | NA | NA | NA | NA | 0.36 |
| athetosis | 1/4 (25.0%) | 1/3 (33.3%) | 0/1 (0.0%) | NA | NA | NA | NA | NA | NA | 0.5 |
| dyskinesia | 18/40 (45.0%) | 12/24 (50.0%) | 3/6 (50.0%) | 2/8 (25.0%) | 1/1 (100.0%) | NA | NA | 0/1 (0.0%) | NA | 0.46 |
| hemifacial spasm | 1/2 (50.0%) | 1/2 (50.0%) | NA | NA | NA | NA | NA | NA | NA | 1 |
| shaking | 38/48 (79.2%) | 25/32 (78.1%) | 3/3 (100.0%) | 4/7 (57.1%) | 2/2 (100.0%) | 2/2 (100.0%) | 1/1 (100.0%) | 1/1 (100.0%) | NA | 0.62 |
| tremor | 174/277 (62.8%) | 79/133 (59.4%) | 25/41 (61.0%) | 36/54 (66.7%) | 20/22 (90.9%)^Mi^ | 5/11 (45.5%) | 6/8 (75.0%) | 3/5 (60.0%) | 0/3 (0.0%)^P^ | **0.03** |
| tremor at rest | 61/138 (44.2%) | 33/76 (43.4%) | 12/23 (52.2%) | 7/20 (35.0%) | 8/10 (80.0%) | 0/1 (0.0%) | 0/4 (0.0%) | 1/3 (33.3%) | 0/1 (0.0%) | 0.12 |
| postural tremor | 52/91 (57.1%) | 31/52 (59.6%) | 9/18 (50.0%) | 6/10 (60.0%) | 2/4 (50.0%) | 0/2 (0.0%) | 3/4 (75.0%) | 1/1 (100.0%) | NA | 0.6 |
| kinetic tremor | 17/35 (48.6%) | 12/20 (60.0%) | 1/7 (14.3%) | 2/4 (50.0%) | 0/1 (0.0%) | NA | 1/2 (50.0%) | 1/1 (100.0%) | NA | 0.27 |
| essential tremor (ET) | 15/18 (83.3%) | 4/7 (57.1%) | 3/3 (100.0%) | 2/2 (100.0%) | 5/5 (100.0%) | NA | 1/1 (100.0%) | NA | NA | 0.23 |
| jaw/chin tremor | 9/23 (39.1%) | 4/17 (23.5%) | 3/3 (100.0%) | 0/1 (0.0%) | NA | NA | 1/1 (100.0%) | 1/1 (100.0%) | NA | 0.04 |
| myoclonus | 11/37 (29.7%) | 9/27 (33.3%) | 1/3 (33.3%) | 1/4 (25.0%) | 0/1 (0.0%) | 0/1 (0.0%) | 0/1 (0.0%) | NA | NA | 0.91 |
| cortical myoclonus | 1/1 (100.0%) | 1/1 (100.0%) | NA | NA | NA | NA | NA | NA | NA | 1 |
| limb myoclonus | 1/3 (33.3%) | 0/2 (0.0%) | NA | NA | NA | NA | NA | 1/1 (100.0%) | NA | 0.08 |
| facial myoclonus | 3/5 (60.0%) | 2/4 (50.0%) | 1/1 (100.0%) | NA | NA | NA | NA | NA | NA | 0.36 |
| finger myoclonus | 0/4 (0.0%) | 0/4 (0.0%) | NA | NA | NA | NA | NA | NA | NA | 1 |
| dystonia | 59/74 (79.7%) | 33/43 (76.7%) | 10/12 (83.3%) | 8/9 (88.9%) | 0/1 (0.0%) | 1/2 (50.0%) | 3/3 (100.0%) | 4/4 (100.0%) | NA | 0.27 |
| retrocollis | 5/6 (83.3%) | 2/3 (66.7%) | 3/3 (100.0%) | NA | NA | NA | NA | NA | NA | 0.27 |
| hand dystonia | 17/21 (81.0%) | 9/12 (75.0%) | 3/3 (100.0%) | 2/2 (100.0%) | 0/1 (0.0%) | NA | 2/2 (100.0%) | 1/1 (100.0%) | NA | 0.27 |
| foot dystonia | 7/7 (100.0%) | 4/4 (100.0%) | 1/1 (100.0%) | 2/2 (100.0%) | NA | NA | NA | NA | NA | 1 |
| upper motor neuron (UMN) signs | 8/13 (61.5%) | 6/9 (66.7%) | NA | 1/2 (50.0%) | NA | 0/1 (0.0%) | NA | NA | 1/1 (100.0%) | 0.49 |
| pseudobulbar palsy | 31/33 (93.9%) | 22/24 (91.7%) | 3/3 (100.0%) | 5/5 (100.0%) | NA | NA | 1/1 (100.0%) | NA | NA | 0.85 |
| inappropriate/pathological laughter | 30/36 (83.3%) | 22/26 (84.6%) | 4/6 (66.7%) | 3/3 (100.0%) | NA | NA | NA | 1/1 (100.0%) | NA | 0.57 |
| inappropriate/pathological crying | 35/50 (70.0%) | 25/33 (75.8%) | 2/6 (33.3%) | 4/5 (80.0%) | NA | 1/1 (100.0%) | 2/2 (100.0%) | 1/2 (50.0%) | 0/1 (0.0%) | 0.2 |
| hyperreflexia | 30/33 (90.9%) | 14/17 (82.4%) | 8/8 (100.0%) | 5/5 (100.0%) | 1/1 (100.0%) | NA | 1/1 (100.0%) | NA | 1/1 (100.0%) | 0.68 |
| positive Hoffmann sign/reflex | 2/8 (25.0%) | 1/4 (25.0%) | NA | 1/2 (50.0%) | 0/1 (0.0%) | NA | NA | NA | 0/1 (0.0%) | 0.72 |
| positive/extensor/upgoing (not absent/flexor/downgoing) Babinski sign/reflex | 16/65 (24.6%) | 12/32 (37.5%) | 3/14 (21.4%) | 0/12 (0.0%) | 0/1 (0.0%) | 0/2 (0.0%) | 0/2 (0.0%) | 1/2 (50.0%) | NA | 0.16 |
| positive/extensor/upgoing (not absent/flexor/downgoing) plantar reflex | 30/150 (20.0%) | 16/76 (21.1%) | 7/30 (23.3%) | 6/32 (18.8%) | 0/3 (0.0%) | 0/4 (0.0%) | 1/3 (33.3%) | 0/1 (0.0%) | 0/1 (0.0%) | 0.9 |
| spasticity | 39/51 (76.5%) | 25/30 (83.3%) | 7/9 (77.8%) | 6/9 (66.7%) | 0/1 (0.0%) | 0/1 (0.0%) | 1/1 (100.0%) | NA | NA | 0.15 |
| lower motor neuron (LMN) signs | 5/12 (41.7%) | 4/8 (50.0%) | NA | 0/2 (0.0%) | NA | 0/1 (0.0%) | 1/1 (100.0%) | NA | NA | 0.29 |
| fasciculations | 6/86 (7.0%) | 3/48 (6.2%)^F^ | 0/10 (0.0%)^F^ | 1/17 (5.9%) | 0/2 (0.0%) | 0/5 (0.0%) | 1/1 (100.0%)^PF,RS^ | 1/2 (50.0%) | 0/1 (0.0%) | **<0.01** |
| tongue atrophy | 6/82 (7.3%) | 3/51 (5.9%) | 1/11 (9.1%) | 2/12 (16.7%) | 0/2 (0.0%) | 0/4 (0.0%) | 0/1 (0.0%) | NA | 0/1 (0.0%) | 0.88 |
| dysarthria | 96/150 (64.0%) | 60/84 (71.4%) | 14/24 (58.3%) | 15/26 (57.7%) | 2/4 (50.0%) | 1/4 (25.0%) | 3/5 (60.0%) | 0/2 (0.0%) | 1/1 (100.0%) | 0.19 |
| dysphasia | 21/30 (70.0%) | 11/14 (78.6%) | 3/5 (60.0%) | 3/5 (60.0%) | 0/1 (0.0%) | 4/5 (80.0%) | NA | NA | NA | 0.47 |
| swallowing difficulty | 140/209 (67.0%) | 86/120 (71.7%) | 17/29 (58.6%) | 26/39 (66.7%) | 4/6 (66.7%) | 1/4 (25.0%) | 5/7 (71.4%) | 1/3 (33.3%) | 0/1 (0.0%) | 0.26 |
| aspiration | 37/56 (66.1%) | 32/42 (76.2%)^RS^ | 1/6 (16.7%)^PF^ | 3/6 (50.0%) | NA | 1/1 (100.0%) | 0/1 (0.0%) | NA | NA | **0.02** |
| PEG/feeding tube insertion | 2/9 (22.2%) | 2/9 (22.2%) | NA | NA | NA | NA | NA | NA | NA | 1 |
| stridor | 3/9 (33.3%) | 2/6 (33.3%) | 1/2 (50.0%) | 0/1 (0.0%) | NA | NA | NA | NA | NA | 0.69 |
| lightheaded/dizziness | 163/211 (77.3%) | 77/107 (72.0%) | 31/36 (86.1%) | 41/50 (82.0%) | 3/4 (75.0%) | 4/4 (100.0%) | 4/5 (80.0%) | 2/3 (66.7%) | 1/2 (50.0%) | 0.52 |
| orthostatic hypotension | 28/56 (50.0%) | 19/39 (48.7%) | 4/8 (50.0%) | 4/6 (66.7%) | 1/2 (50.0%) | 0/1 (0.0%) | NA | NA | NA | 0.79 |
| urinary dysfunction | 117/177 (66.1%) | 67/96 (69.8%) | 17/29 (58.6%) | 15/27 (55.6%) | 5/7 (71.4%) | 5/8 (62.5%) | 5/6 (83.3%) | 2/2 (100.0%) | 1/2 (50.0%) | 0.68 |
| urinary incontinence | 99/151 (65.6%) | 52/79 (65.8%) | 12/22 (54.5%) | 22/29 (75.9%) | 5/7 (71.4%) | 6/9 (66.7%) | 2/4 (50.0%) | NA | 0/1 (0.0%) | 0.54 |
| urinary catheter | 13/15 (86.7%) | 8/9 (88.9%) | 1/1 (100.0%) | 1/2 (50.0%) | 1/1 (100.0%) | 1/1 (100.0%) | 1/1 (100.0%) | NA | NA | 0.7 |
| urinary tract infection (UTI) | 19/22 (86.4%) | 11/13 (84.6%) | 4/4 (100.0%) | 2/3 (66.7%) | 1/1 (100.0%) | NA | 1/1 (100.0%) | NA | NA | 0.74 |
| erectile dysfunction (ED) | 27/28 (96.4%) | 17/18 (94.4%) | 3/3 (100.0%) | 4/4 (100.0%) | 1/1 (100.0%) | 2/2 (100.0%) | NA | NA | NA | 0.97 |
| abnormal sweat | 17/50 (34.0%) | 12/33 (36.4%) | 1/8 (12.5%) | 3/4 (75.0%) | 0/2 (0.0%) | 0/1 (0.0%) | 1/2 (50.0%) | NA | NA | 0.26 |
| cold discolored hands/feet | 8/9 (88.9%) | 6/7 (85.7%) | 2/2 (100.0%) | NA | NA | NA | NA | NA | NA | 0.57 |
| improvement with l-dopa, levodopa or sinemet | 65/112 (58.0%) | 44/77 (57.1%) | 11/17 (64.7%) | 7/12 (58.3%) | 1/3 (33.3%) | 1/2 (50.0%) | NA | 1/1 (100.0%) | NA | 0.87 |

PSP, progressive supranuclear palsy; PSP-RS, PSP-Richardson’s syndrome; PSP-PGF, PSP-progressive gait freezing; PSP-P, PSP-parkinsonism; PSP-SL, PSP-speech/language disorders; PSP-CBS, PSP-corticobasal syndrome; CBS, Corticobasal syndrome; PSP-F, PSP- frontal variant; PSP-C, PSP-cerebellar variant; O1-2, level 1 or level 2 of ocular motor dysfunction; P1-2, level 1 or level 2 of postural instability; NA, not available

The number of cases with a particular syndrome/symptom/sign within three years after onset are evaluated and are presented as “number of cases present” / “number of cases evaluated by three years after onset” (%). Overall statistical differences for each variable are evaluated using the chi-square test, followed by the pairwise chi-square test with Holm’s correction for those with significant differences. *P* value < 0.05 is considered statistically significant. Superscripts 1 to 5 mean *P* < 0.05 vs. subtype 1 to 5, respectively
